# Estimating typhoid incidence from community-based serosurveys: A multicohort study in Bangladesh, Nepal, Pakistan and Ghana

**DOI:** 10.1101/2021.10.20.21265277

**Authors:** Kristen Aiemjoy, Jessica C. Seidman, Senjuti Saha, Sira Jam Munira, Mohammad Saiful Islam Sajib, Syed Muktadir Al Sium, Anik Sarkar, Nusrat Alam, Farha Nusrat Jahan, Md. Shakiul Kabir, Dipesh Tamrakar, Krista Vaidya, Rajeev Shrestha, Jivan Shakya, Nishan Katuwal, Sony Shrestha, Mohammad Tahir Yousafzai, Junaid Iqbal, Irum Fatima Dehraj, Yasmin Ladak, Noshi Maria, Mehreen Adnan, Sadaf Pervaiz, Alice S. Carter, Ashley T. Longley, Clare Fraser, Edward T. Ryan, Ariana Nodoushani, Alessio Fasano, Maureen Leonard, Victoria Kenyon, Isaac I. Bogoch, Hyon Jin Jeon, Andrea Haselbeck, Se Eun Park, Raphael Zellweger, Florian Marks, Ellis Owusu-Dabo, Yaw Adu-Sarkodie, Michael Owusu, Peter Teunis, Stephen P. Luby, Denise O. Garrett, Farah Naz Qamar, Samir K. Saha, Richelle C. Charles, Jason R. Andrews

## Abstract

**Background:** The incidence of enteric fever, an invasive bacterial infection caused by typhoidal *Salmonellae*, is largely unknown in regions lacking blood culture surveillance. New serologic markers have proven accurate in diagnosing enteric fever, but whether they could be used to reliably estimate population-level incidence is unknown.

**Methods:** We collected longitudinal blood samples from blood culture-confirmed enteric fever cases enrolled from surveillance studies in Bangladesh, Nepal, Pakistan, and Ghana and conducted cross-sectional serosurveys in the catchment areas of each surveillance site. We used ELISAs to measure quantitative IgA and IgG antibody responses to Hemolysin E (HlyE) and *S*. Typhi lipopolysaccharide (LPS). We used Bayesian hierarchical models to fit two-phase power-function decay models to the longitudinal antibody responses among enteric fever cases and used the joint distributions of the peak antibody titers and decay rate to estimate population-level incidence rates from cross-sectional serosurveys.

**Findings:** The longitudinal antibody kinetics for all antigen-isotypes were similar across countries and did not vary by clinical severity. The seroincidence of typhoidal *Salmonella* infection among children <5 years ranged between 58.5 per 100 person-years (95% CI: 42.1 - 81.4) in Dhaka, Bangladesh to 6.6 (95% CI: 4.3-9.9) in Kavrepalanchok, Nepal, and followed the same rank order as clinical incidence estimates.

**Interpretation:** The approach described here has the potential to expand the geographic scope of typhoidal *Salmonella* surveillance and generate incidence estimates that are comparable across geographic regions and time.

**Funding:** This work was supported by the Bill and Melinda Gates Foundation (INV-000572).

**Research in context:** *Evidence before this study:* Previous studies have identified serologic responses to two antigens (Hemolysin E [HlyE] and *Salmonella* lipopolysaccharide [LPS]) as promising diagnostic markers of acute typhoidal *Salmonella* infection. We reviewed the evidence for seroepidemiology tools for enteric fever available as of November 01, 2021, by searching the National Library of Medicine article database and *medRxiv* for preprint publications, published in English, using the terms “enteric fever”, “typhoid fever”, “*Salmonella* Typhi”, “*Salmonella* Paratyphi”, “typhoidal *Salmonella*”, “Hemolysin E”, “*Salmonella* lipopolysaccharide”, “seroconversion”, “serosurveillance”, “seroepidemiology”, “seroprevalence” and “seropositivity.” We found no studies using HlyE or LPS as markers to measure the incidence or prevalence of enteric fever in a population. Anti-Vi IgG responses were used as a marker of population seroprevalence in cross-sectional studies conducted in South Africa, Fiji, and Nepal, but were not used to calculate population-based incidence estimates.

*Added value of this study:* We developed and validated a method to estimate typhoidal *Salmonella* incidence in cross-sectional population samples using antibody responses measured from dried blood spots. First, using longitudinal dried blood spots collected from over 1400 blood culture-confirmed cases in four countries, we modeled the longitudinal dynamics of antibody responses for up to two years following infection, accounting for heterogeneity in antibody responses and age-dependence. We found that longitudinal antibody responses were highly consistent across four countries on two continents and did not differ by clinical severity. We then used these antibody kinetic parameters to estimate incidence in population-based samples in six communities across the four countries, where concomitant population-based incidence was measured using blood cultures. Seroincidence estimates were much higher than blood-culture-based case estimates across all six sites, suggestive of a high incidence of asymptomatic or unrecognized infections. Still, the rank order of seroincidence and culture-based incidence rates were the same, with the highest rates in Bangladesh and lowest in Ghana.

*Implications of all the available evidence:* Many at-risk low- and middle-income countries lack data on typhoid incidence needed to inform and evaluate vaccine introduction. Even in countries where incidence estimates are available, data are typically geographically and temporally sparse due to the resources necessary to initiate and sustain blood culture surveillance. We found that typhoidal *Salmonella* infection incidence can be estimated from community-based serosurveys using dried blood spots, representing an efficient and scalable approach for generating the typhoid burden data needed to inform typhoid control programs in resource-constrained settings.

## INTRODUCTION

Enteric fever, an invasive infection caused by *Salmonella enterica* subspecies *enterica* serovars Typhi and Paratyphi A, B, or C, is a significant cause of preventable morbidity and mortality in low- and middle-income countries (LMICs) (1). Enteric fever incidence is typically ascertained using clinical surveillance, where blood culture-positive cases are tallied and reported relative to a catchment area population. Blood culture requires considerable laboratory infrastructure not widely accessible in many LMICs. Another limitation is that only individuals seeking care at surveillance sites are captured, whereas many patients receive treatment from healthcare providers outside the reach of traditional surveillance systems. Even when available, the estimated sensitivity of blood culture is only 60% (2). Consequently, blood culture surveillance covers a small proportion of at-risk populations worldwide and underestimates the burden of disease.

Population-based serologic surveillance (serosurveillance) can estimate infection transmission in settings lacking facility-based surveillance. Cross-sectional serosurveillance has been helpful for rapidly characterizing transmission of COVID-19, pertussis, dengue, and other diseases (3–7). However, serosurveillance for typhoidal *Salmonella* has been limited by the lack of sensitive and specific serologic markers. Serologic responses to the most widely used antigen to date, virulence (Vi) capsular polysaccharide, have poor diagnostic performance during acute infection and cannot distinguish between natural infection and Vi-based vaccination (8). Immune responses to Hemolysin E (HlyE) and *S*. Typhi lipopolysaccharide (LPS) demonstrate diagnostic accuracy for acute enteric fever (9–11) but have not been evaluated as tools to measure population-level seroincidence. HlyE, a cytotoxic pore-forming toxin that invades epithelial cells (12–14), is present in *S*. Typhi and Paratyphi A but rarely found in other serovars (15). LPS is a major component of the outer membrane of Gram-negative organisms and a potent inducer of innate immunity (16).

Here, we model longitudinal immune responses to HlyE and LPS among blood culture-confirmed enteric fever patients enrolled from two multi-year, hospital-based, enteric fever surveillance studies: The Surveillance for Enteric Fever in Asia Project (SEAP) in Bangladesh, Nepal, and Pakistan, (17) and the Severe Typhoid in Africa (SETA) surveillance study in Ghana (18). We then use the longitudinal antibody dynamics to estimate typhoidal *Salmonella* seroincidence from cross-sectional population data and compare seroincidence rates to clinical incidence rates in the same catchment areas (17,18).

## METHODS

### Overview

We enrolled blood culture-confirmed enteric fever cases and collected longitudinal blood samples for up to two years after infection. Concurrently, we conducted population-based serosurveys in the catchment areas of each clinical surveillance site. We characterized the dynamics of antibody responses to HlyE and LPS among cases, then applied a model using these kinetics to cross-sectional data to estimate the population-level seroincidence of infection.

### Study populations and enrollment

#### Enteric fever cases

All blood culture-positive enteric fever cases enrolled through SEAP were eligible to participate in the ancillary serologic study. Patients were enrolled from five hospitals: Dhaka Shishu Hospital in Dhaka, Bangladesh; Kathmandu Medical College and Teaching Hospital (KMC) in Kathmandu and Dhulikhel Hospital, Kathmandu University Hospital (DH) in Kavre District, Nepal; and Aga Khan University Hospital (AKU) and Kharadar General Hospital (KGH) in Karachi, Pakistan; and a network of laboratory facilities in all three countries between 2016 and 2021. During the enrollment visit, information on demographics, symptom history and typhoid vaccination status were collected using a structured questionnaire; SEAP enrollment criteria and methods are detailed elsewhere (17). For prospective cases, we collected plasma at SEAP enrollment and capillary blood collected on filter paper (i.e., dried blood spot; DBS) at 28 days, 3, 6, 12, and 18 months post-enrollment. We also enrolled cases retrospectively and collected DBS at enrollment and subsequent scheduled visits following the above timeline. For SETA-Ghana, blood culture-confirmed enteric fever patients were enrolled from the Agogo Presbyterian Hospital and the Komfo Anokye Teaching Hospital in Agogo. SETA study design and methodology are published elsewhere (18). Plasma collected at days 3-7, 28-30, 90, 180, 270, and 360 were included in our analysis. Baseline plasma samples were also collected from 17 invasive non-typhoidal *Salmonella* (iNTS) cases.

#### Population-based samples

We enrolled geographically random, population-based samples of individuals aged 0 to 25 years from the catchment areas of SEAP hospitals between 2019 and 2021 (Figure S1). We focused on our serosurveys on individuals under the age of 26, because most patients (whose data was used to parameterize the longitudinal kinetic model) were under 26 years of age (86.5%; 1228/1420). We randomly selected sampling grid clusters within catchment areas, enumerated all households residing in each cluster, and then randomly selected an age-stratified sample (0-4, 5-9, 10-15, and 16-25 years). As we sought to enroll a broad representative sample of the population, there were no exclusion criteria other than age and residence in the study catchment area for participating in the cross-sectional survey. For the SETA-Ghana site, we collected plasma from up to four neighborhood controls, matched by age, sex, and enrollment site between 2016 and 2018 (Figure S1).

### Laboratory Methods

Capillary blood samples were collected on TropBioTM filter papers (Cellabs Pty Ltd., Brookvale, New South Wales, Australia), air-dried for ≥2 hours at room temperature, stored with desiccant in individual plastic bags at −20 □C until processing. Plasma was stored at −70 □C until processing. Study laboratories analyzed SEAP samples in each country; SETA and North American samples were analyzed at MGH. We used kinetic enzyme-linked immunosorbent assays (ELISAs) to quantify antibody levels in plasma and eluted DBS samples. Details on the ELISA methods are provided in the **appendix** along with a comparison of antibody responses measured from DBS and Plasma (Figure S2).

### Statistical Methods

To describe the antibody kinetics of each antigen-isotype following typhoidal *Salmonella* infection, we fit two-phase models with an exponential rise, peak, and then power-function decay episode (19,20). We fit the models using a Bayesian hierarchical framework obtaining predictive posterior samples using Markov chain Monte Carlo (MCMC) sampling for baseline (y0) and peak antibody responses (y1), time to peak (t1), and decay rate (α) and decay shape (r) (20). The models were run in JAGS, using the rjags package (21). We fit the longitudinal antibody response models in three age strata: <5 years, 5-15 years, >15 years chosen to compare the seroincidence estimates to clinical incidence estimates from the same catchment areas. We report the median and 95% credible interval (CrI) from each posterior distribution.

When following individuals over time, reinfections are possible and increase in likelihood with longer follow-up. We defined suspected reinfections as the occurrence of a ≥3-times increase in antibody response in individuals between visits ≥3 months from fever onset for ≥2 antigen-isotype combinations unless the absolute value of the difference between measurements was <1 ELISA unit. The 3-times threshold was derived by calculating the median times change from baseline to 28 days among blood culture-confirmed cases. Additional details on how we defined reinfections are available in the appendix. Observations including and after the suspected reinfection event were excluded from the longitudinal decay parameter estimation.

To estimate seroincidence, we created a likelihood function for the observed cross-sectional population data based on the longitudinal kinetics following infection (22). We assumed that incident infections in the study sample occurred as a Poisson process with rate (*λ*) and generate maximum likelihood profiles for *λ* with each antigen and isotype separately and also jointly estimated *λ* by combining their likelihood functions (23). We estimated age-stratified incidence rates in the population using the age-specific antibody response parameters. We accounted for two sources of noise in the observed serologic responses: measurement noise of the assay (described by the coefficient of variation [CV] across replicates) and biologic noise (measured as background response to the antigen-isotype among never-exposed, negative controls) as detailed in (22). Details on how we specified the measurement and biologic noise parameters are provided in the appendix. We used mixed-effect models to adjust the standard errors for clustering by sampling unit for the population-based serosurvey).

We compared our population-based seroincidence estimates to clinical blood culture-based incidence estimates derived from the same catchment area populations. We included both crude incidence (culture-confirmed cases divided by the catchment population and observation time) and adjusted incidence estimates (accounting for blood culture sensitivity and the proportion of patients with typhoid-like illness who had blood culture at a surveillance site), following previously described methods (24). We estimated the seroincidence rates among children ages 2 to <5 compared with the clinical incidence estimates reported for this age category. We focused the comparisons among young children, as the seroincidence will reflect recent infections during the same period as the clinical incidence study. While clinical incidence estimates were available for some catchment areas for children under two years, we had an insufficient number of young children under 2 in the population samples to estimate seroincidence in this age stratum.

### Ethical considerations

We obtained written informed consent from all eligible participants and the parents/guardians of participants aged <18 years before collecting blood samples and completing the questionnaire; we obtained written assent from children aged 15 to 17 years in Bangladesh, Nepal, and Pakistan and aged 12 to 17 in Ghana. Institutional Review Boards in the United States (Centers for Disease Control and Prevention; Stanford University Institutional Review Board), Bangladesh (Bangladesh Institute of Child Health Ethical Review Committee), Nepal (Nepal Health Research Council Ethical Review Board), Pakistan (AKU Ethical Review Committee and Pakistan National Bioethics Committee), Korea (International Vaccine Institute IRB), Belgium (Institute of Tropical Medicine Antwerp Institutional Review Board) and Ghana (Komfo Anokye Teaching Hospital, Committee on Human Research, Publication and Ethics) approved the study forms and protocols.

## RESULTS

We enrolled a longitudinal cohort of 1420 blood culture-confirmed enteric fever cases between 2017 and early 2021 (407 from Bangladesh, 543 from Nepal, 399 from Pakistan, and 71 from Ghana) from the SEAP and SETA studies (Table 1). We followed cases for a median of 382 days after fever onset (interquartile range 94 – 696 days) and collected and analyzed 4126 longitudinal blood samples (Figure S3). Median responses at 1 and 6 months after fever onset were comparable across sites and higher than the median values for the serosurvey participants at each location. (Figure S4). Correlation between antigen-isotype responses was higher among younger cases and at earlier time points (closer to infection) (Figure S5).

**Table 1.** Demographic, clinical and sampling characteristics of culture-confirmed enteric fever cases included in the longitudinal kinetic analysis, by country.

|  | **Bangladesh**<br>**(N=407)** | **Nepal**<br>**(N=543)** | **Pakistan**<br>**(N=399)** | **Ghana**<br>**(N=71)** | **Total**<br>**(N=1420)** |
| --- | --- | --- | --- | --- | --- |
| **Age in years** |  |  |  |  |  |
| Median | 5.4 | 20.9 | 5.3 | 8.0 | 9.0 |
| Q1, Q3 | 3.2, 8.0 | 15.7, 26.4 | 3.0, 12.0 | 6.0, 12.0 | 4.5, 19.7 |
| **Age strata** |  |  |  |  |  |
| <5 | 179 (44.0%) | 12 (2.2%) | 181 (45.4%) | 11 (15.5%) | 383 (27.0%) |
| 5-15 | 227 (55.8%) | 127 (23.4%) | 152 (38.1%) | 52 (73.2%) | 558 (39.3%) |
| 16+ | 1 (0.2%) | 404 (74.4%) | 66 (16.5%) | 8 (11.3%) | 479 (33.7%) |
| **Sex** |  |  |  |  |  |
| Male | 213 (52.3%) | 302 (55.6%) | 228 (57.1%) | 41 (58.6%) | 784 (55.3%) |
| Female | 194 (47.7%) | 241 (44.4%) | 171 (42.9%) | 29 (41.4%) | 635 (44.7%) |
| **Number of sera samples collected per participant** |  |  |  |  |  |
| Median | 2.0 | 2.0 | 3.0 | 4.0 | 3.0 |
| Q1, Q3 | 2.0, 3.0 | 2.0, 4.0 | 2.0, 5.0 | 1.5, 5.0 | 2.0, 4.0 |
| **Study enrollment time, in days** |  |  |  |  |  |
| Median | 184.0 | 366.0 | 699.0 | 270.0 | 382.0 |
| Q1, Q3 | 28.0, 540.5 | 160.0, 595.5 | 367.5, 712.0 | 14.5, 365.0 | 94.0, 696.0 |
| **Reported days of fever at presentation** |  |  |  |  |  |
| Median | 4.0 | 4.0 | 6.0 | 6.0 | 4.0 |
| Q1, Q3 | 3.0, 6.0 | 3.0, 6.0 | 4.0, 9.0 | 3.0, 7.0 | 3.0, 7.0 |
| **Hospitalized** |  |  |  |  |  |
| No | 326 (80.1%) | 372 (70.5%) | 210 (52.8%) | 31 (48.4%) | 939 (67.2%) |
| Yes | 81 (19.9%) | 156 (29.5%) | 188 (47.2%) | 33 (51.6%) | 458 (32.8%) |
| **Serovar** |  |  |  |  |  |
| S. Paratyphi A | 56 (13.8%) | 86 (15.8%) | 11 (2.8%) | 0 (0.0%) | 153 (10.8%) |
| S. Typhi | 351 (86.2%) | 457 (84.2%) | 388 (97.2%) | 71 (100.0%) | 1267 (89.2%) |
| **Vaccine status** |  |  |  |  |  |
| TCV | 1 (0.2%) | 1 (0.2%) | 4 (1.0%) | 0 (0.0%) | 6 (0.4%) |
| Other Typhoid Vaccine | 1 (0.2%) | 1 (0.2%) | 8 (2.1%) | 0 (0.0%) | 10 (0.7%) |
| Not Vaccinated | 405 (99.5%) | 541 (99.6%) | 378 (96.9%) | 71 (100.0%) | 1395 (98.9%) |

Thirty-seven participants had a three-times or higher increase in antibody responses meeting the definition of suspected reinfection. The reinfection incidence was 5.2 (95% CI 1.8 - 8.5) in Bangladesh, 4.9 (95% CI: 4.0 - 6.8) in Pakistan, 0.7 (96% CI: 0.0 - 1.6) in Nepal, and 5.6 (95% CI: 0 - 13.3) in Ghana per 100-person years. The median time to detection of suspected reinfection was 13.8 months following fever onset (IQR: 10.3-18.4 months).

Antibody responses to HlyE and LPS reached peak levels within three weeks of fever onset (Figure 1A). Peak HlyE antibody responses increased with age. The rate of antibody decay decreased with age across all antigen-isotype combinations (Figure 1B; Table S1). The overall decay rate for HlyE IgA was slightly faster than for HlyE IgG. The shape parameters for all antigen-isotypes deviate from 1, indicating non-exponential decay (Table S1). Median responses remained elevated above baseline levels for 29 months for HlyE IgG, 14 months for LPS IgG, 11 months for HlyE IgA, and 3 months for LPS IgA.

**Fig. 1.**
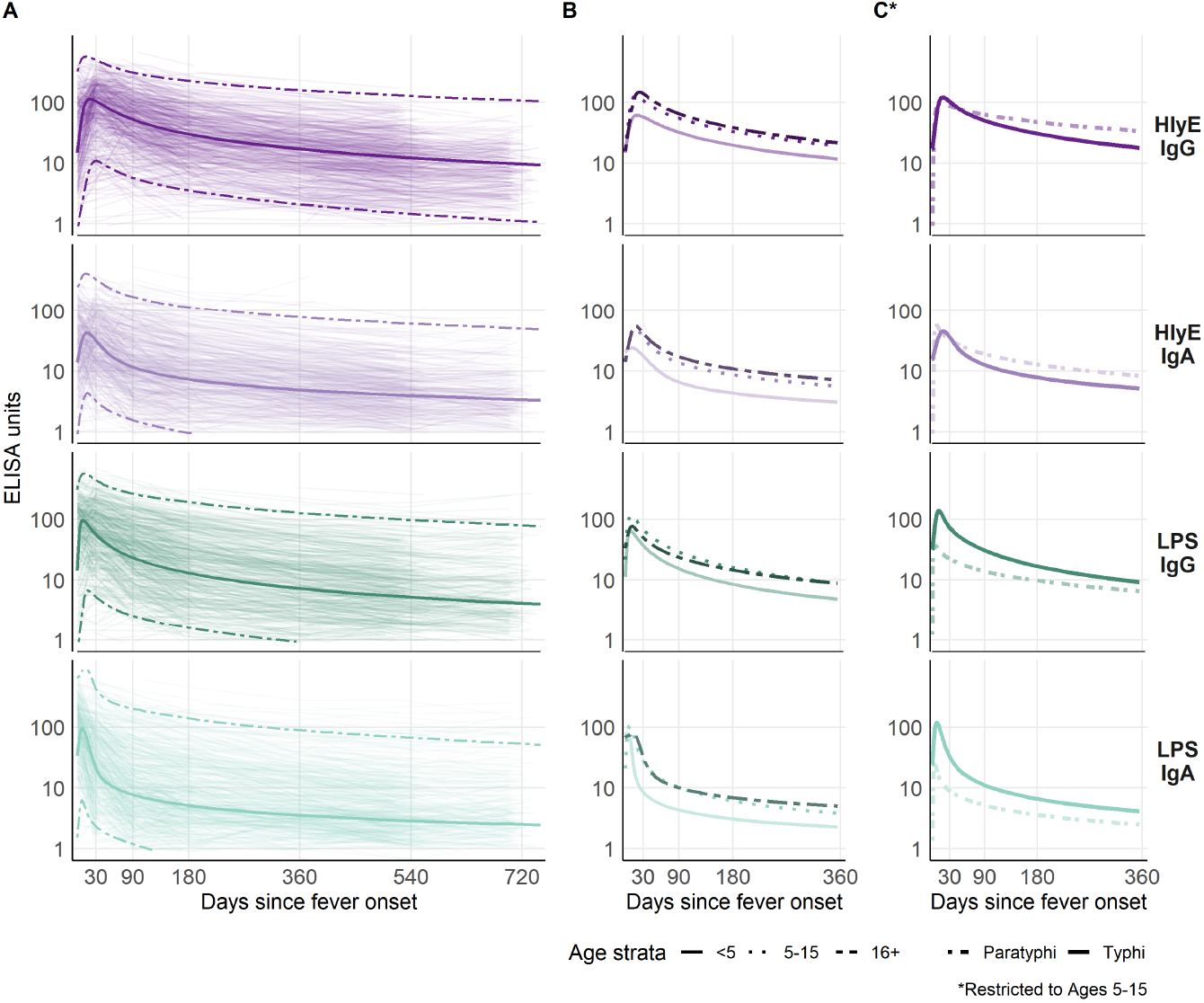
Longitudinal antibody dynamics among blood culture-confirmed enteric fever cases. Longitudinal antibody dynamics were fit to ELISA-measured antibody responses using Bayesian hierarchical models. (A) The light-colored lines are the observed individual antibody concentrations; each line indicates one patient. The dark solid and dotted lines indicate the median and 95% credible intervals for the model-fitted antibody decay concentrations. The median model-fitted antibody trajectories are shown for each age strata (B) and comparing *Salmonella* Typhi to *Salmonella* Paratyphi A cases (C). Panel (C) restricts to cases 5-15 years old because *S*. Paratyphi A cases were older than *S*. Typhi cases.

The model fitted antibody trajectories were similar across all four study sites (Figure 2A). The distributions for peak antibody responses were nearly identical across countries (Figure 2B), and the differences between distributions all centered near 0 for all antigen-isotype combinations. There was some inter-country variation in the decay rate, with cases in Bangladesh having a slower decay rate than the other sites (Figure 2C).

**Fig. 2.**
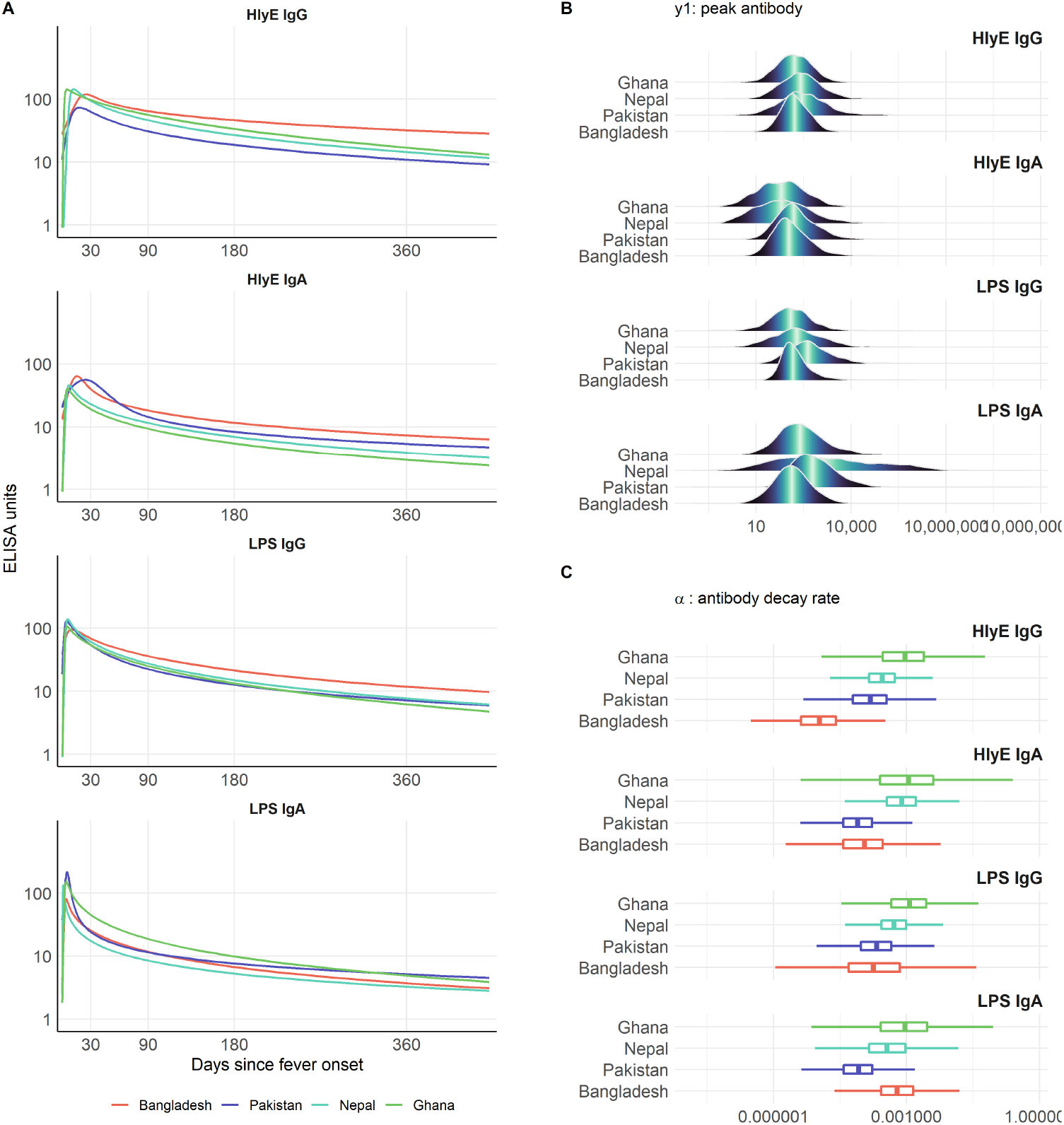
Antibody kinetics, peak response and decay rate among blood culture-confirmed enteric fever cases by study country. (A) Longitudinal antibody decay profiles fit to ELISA-measured antibody responses in each study country. (B) The distribution of model-predicted peak antibody responses in each study country. (C) Boxplots and distributions of model-predicted antibody decay rates across study countries. All comparisons are among children 5 to 15 years to account for the different age-distribution of cases across countries.

*S*. Paratyphi A was responsible for 10.8% (153/1136) of enteric fever cases overall: 13.8% (56/407) in Bangladesh, 15.8% (86/543) in Nepal, 2.8% (11/399) in Pakistan and 0% (0/71) in Ghana (Table 1). Compared with *S*. Typhi cases, *S*. Paratyphi A cases had similar HlyE IgG and IgA antibody trajectories; however, peak responses for both LPS IgA and IgG were lower (Figure 1C). We also compared antibody responses among enteric fever cases who were hospitalized (32.8%; (458/1420) and not hospitalized, and found similar antibody trajectories for all antigen-isotype combinations (Figure S6).

In parallel to the longitudinal cohort, we enrolled and analyzed DBS from 1740 children and young adults in cross-sectional population-based serosurveys: 401 in Dhaka, Bangladesh; 353 in Kathmandu and 481 in Kavre, Nepal, 294 from the AKU catchment area, and 200 from the KGH catchment area in Karachi, Pakistan, and 79 in Agogo, Ghana (Figure S7; Table 2). Median antibody responses for all antigen-isotypes increased with age and were highest in Dhaka, Bangladesh, and lowest in Kavre, Nepal, and Agogo, Ghana (Figure S8). Compared to the antibody responses observed in cases within six months of fever onset, median values for the serosurvey participants were lower for all antigen-isotypes at each site (Figure S4).

**Table 2.** Demographic and sampling characteristics of the population-based cross-sectional serosurvey participants by country

|  | **Bangladesh: Dhaka**<br>**(N=401)** | **Nepal: Kathmandu**<br>**(N=353)** | **Nepal: Kavre**<br>**(N=481)** | **Pakistan: AKU**<br>**(N=294)** | **Pakistan: KGH**<br>**(N=200)** | **Ghana: Agogo**<br>**(N=79)** |
| --- | --- | --- | --- | --- | --- | --- |
| **Age in years** |  |  |  |  |  |  |
| Median | 9.2 | 12.0 | 10.2 | 10.0 | 8.0 | 6.0 |
| Q1, Q3 | 4.9, 14.0 | 5.8, 17.8 | 5.1, 15.7 | 4.9, 16.0 | 4.9, 13.7 | 5.0, 9.0 |
| **Age strata** |  |  |  |  |  |  |
| <5 | 101 (25.2%) | 73 (20.7%) | 113 (23.5%) | 75 (25.5%) | 51 (25.5%) | 18 (22.8%) |
| 5-15 | 256 (63.8%) | 169 (47.9%) | 249 (51.8%) | 143 (48.6%) | 118 (59.0%) | 59 (74.7%) |
| 16-25 | 44 (11.0%) | 111 (31.4%) | 119 (24.7%) | 76 (25.9%) | 31 (15.5%) | 2 (2.5%) |
| **Gender** |  |  |  |  |  |  |
| Male | 183 (45.6%) | 184 (52.1%) | 248 (51.6%) | 135 (45.9%) | 97 (48.5%) | 47 (59.5%) |
| Female | 218 (54.4%) | 169 (47.9%) | 233 (48.4%) | 159 (54.1%) | 103 (51.5%) | 32 (40.5%) |
| **Vaccine Status** |  |  |  |  |  |  |
| TCV | 13 (3.2%) | 0 (0.0%) | 0 (0.0%) | 127 (43.2%) | 85 (42.5%) | 0 (0.0%) |
| Other Typhoid<br>Vaccine | 17 (4.2%) | 3 (0.9%) | 1 (0.2%) | 3 (1.0%) | 1 (0.5%) | 0 (0.0%) |
| Not Vaccinated | 371 (92.5%) | 348 (99.1%) | 480 (99.8%) | 164 (55.8%) | 114 (57.0%) | 79 (100.0%) |

We used only HlyE IgG and IgA to estimate seroincidence because 1) LPS antibody responses were lower among *S*. Paratyphi A cases (Figure 2C) and 2) we identified elevated LPS antibody responses among invasive non-typhoidal Salmonellosis (iNTS) patients *(20, 21)* (Figure S9), suggesting possible cross-reactivity. The seroincidence rate of enteric fever per 100 person-years was highest in Dhaka, Bangladesh (41.2, 95% CI: 34.0-50.1), followed by Karachi, Pakistan (17.6, 95% CI: 13.7-22.6 in KHG; 10.5, 95% CI: 8.5-13.0 in AKU), Nepal (6.6, 95% CI: 5.4-8.1 in Kathmandu; 5.8, 95% CI: 4.8-7.1 in Kavre) and Agogo, Ghana (5.5, 95% CI: 4.1-7.35) (Figure 3). In Dhaka, Agogo, and the KGH and AKU catchment areas, the seroincidence was higher in the youngest age group (<5 years). In Kathmandu and kavre, the seroincidence was similar across age strata. Seroincidence estimates using anti-HlyE and anti-LPS IgA and IgG are presented in the **appendix** (Table S2). The differences in incidence estimates using overall versus country-specific longitudinal parameters (peak, decay rate, and decay shape) are presented in Figure S10.

**Fig. 3.**
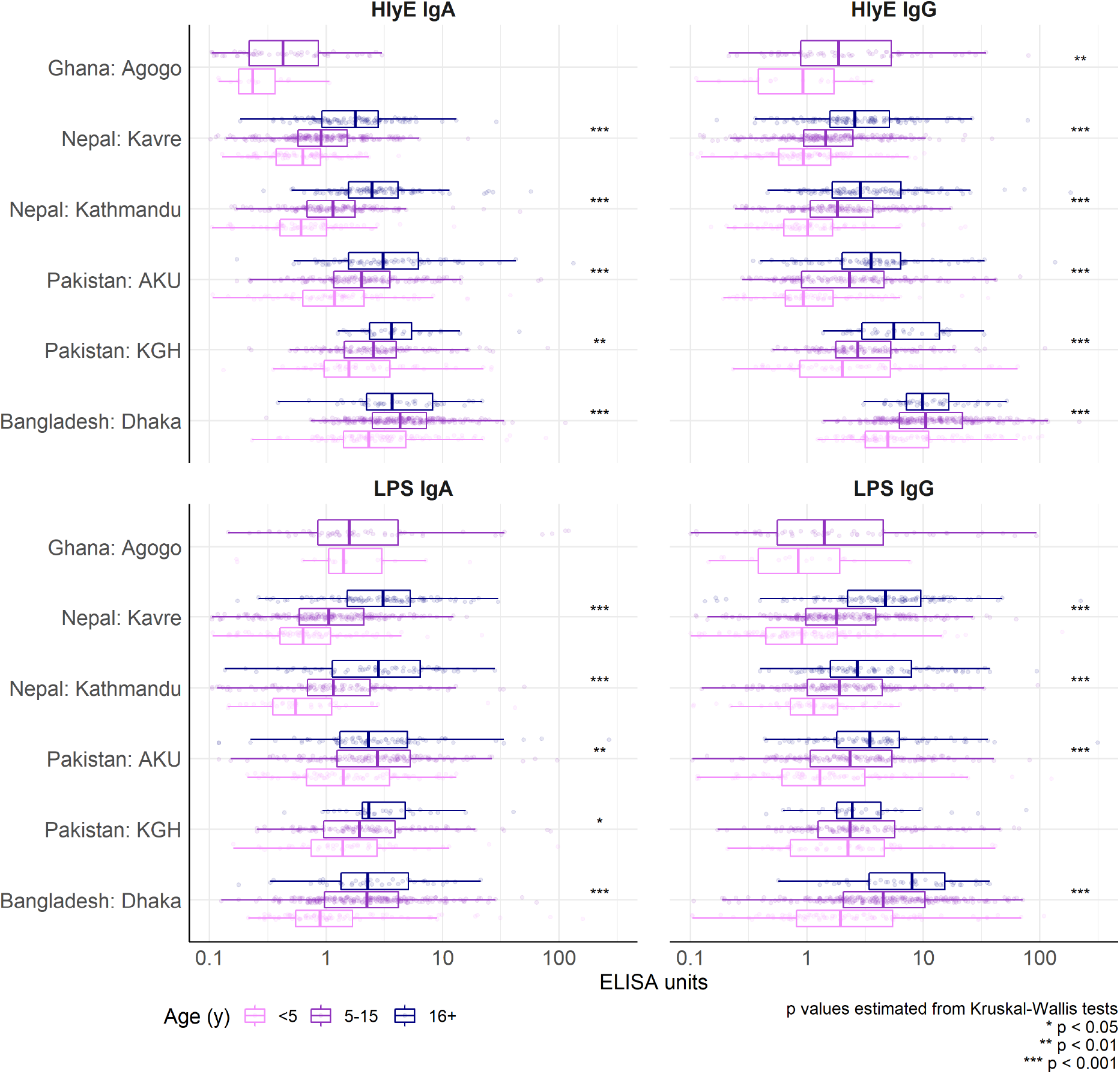
Age trends in serologic responses to HlyE and LPS in cross-sectional serosurveys from six communities. Anti-HlyE and LPS IgA and IgG responses are compared across age strata and catchment area populations. Boxes indicate the median and interquartile range. Antibody responses were measured using kinetic enzyme-linked immunosorbent assays (ELISAs). Asterisks denote levels of statistical significance for differences across age strata using the Kruskal-Wallis test.

Among young children 2-4 years old, the seroincidence estimates followed the same rank order as the clinical incidence estimates for each catchment area. Dhaka, Bangladesh, had the highest seroincidence, followed by KGH and AKU in Karachi, Pakistan then Kathmandu and Kavre, in Nepal (Figure 4). Seroincidence estimates were between 16 and 32 times higher than care-seeking adjusted clinical incidence rates (incidence ratio: 32.3 in Bangladesh; 28.9 in KGH; 26.1 in AKU; 36.7 in Kavre; and 15.9 in Kathmandu).

**Fig. 4.**
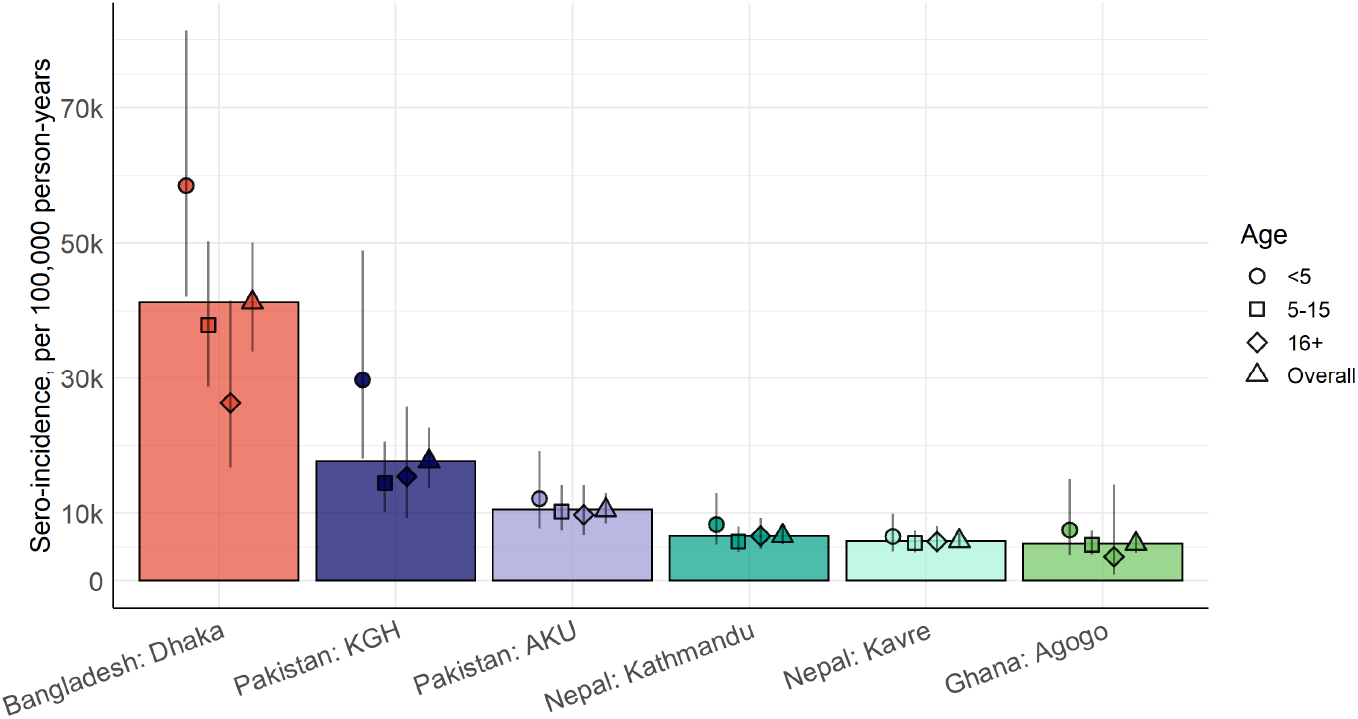
Estimated seroincidence of typhoidal *Salmonella* by study community and age group. Age groups are denoted by point shapes for the median, with lines indicating the 95% confidence intervals. Boxes reflect the height of the median estimate for the overall population-based serosurvey.

**Fig. 5.**
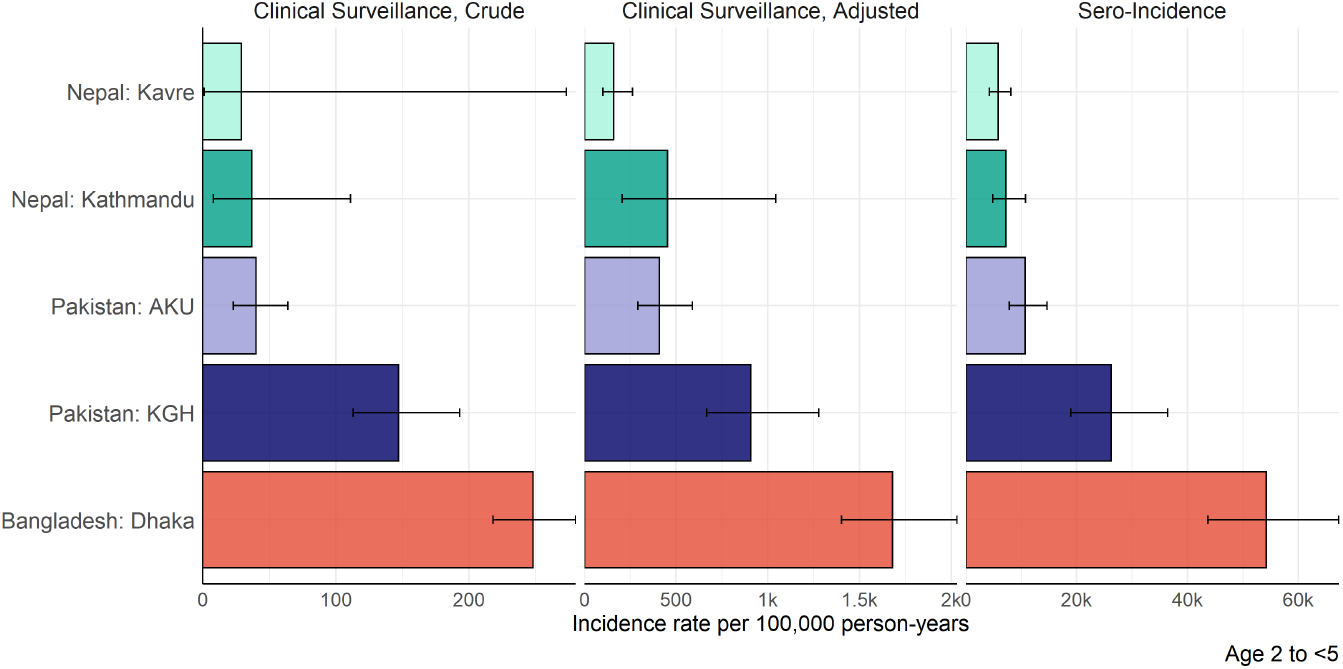
Comparison of estimates for crude and adjusted clinical enteric fever incidence with typhoidal *Salmonella* seroincidence. Crude clinical incidence reflects culture-confirmed *S*. Typhi and *S*. Paratyphi A cases divided by the catchment population and time, while adjusted incidence accounts for imperfect sensitivity of blood culture and the proportion of acute febrile illnesses captured by the surveillance system. The horizontal axis indicates incidence, and scale differs for type of estimate. Estimates are shown for children 2-4 years of age, for the serological estimates to coincide with the age stratification of clinical surveillance.

**Fig. 6.**
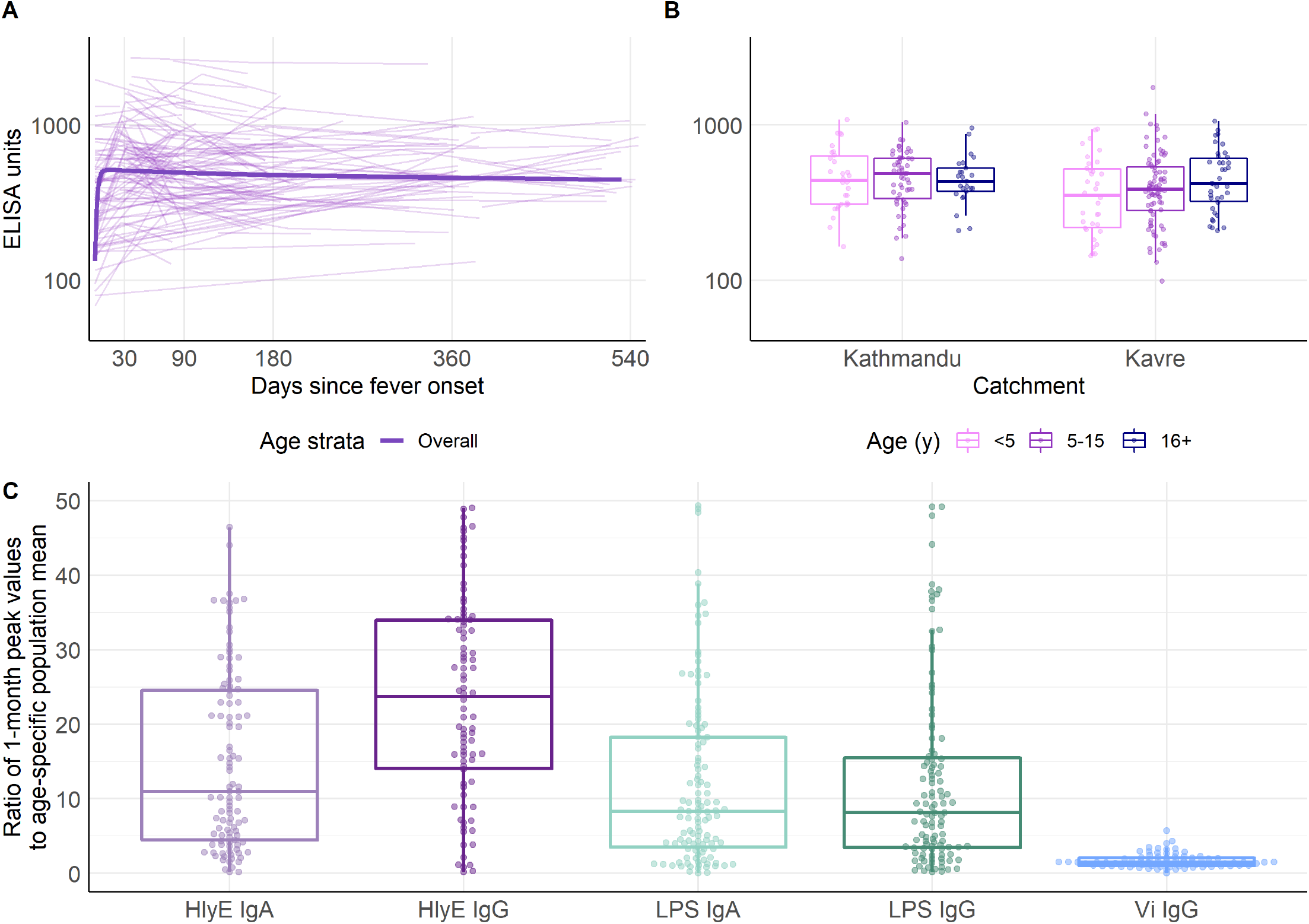
Kinetics and age-dependent anti-Vi IgG antibody responses for Vi IgG. Data are from the two study communities in Nepal. (A) Anti-Vi IgG responses following fever onset among culture-confirmed *S*. Typhi cases. (B) Anti-Vi IgG responses by age from the cross-sectional population-based serosurveys in two communities. (C) Ratio of antibody responses among culture-confirmed typhoid cases at 1 month following fever onset, compared with the mean of the population-based serosurvey participants.

In a sub-analysis of samples from blood culture-positive cases in Nepal, we found that after a modest rise, anti-Vi antibody responses plateaued with no subsequent decay (rate = 0) (Figure S11 Table S1). Median anti-Vi IgG levels remained elevated above baseline for at least 32 months and did not vary significantly by age (Figure 11B, p > 0.05). The ratio of 28-day responses among cases compared with the population-level mean for each antigen-isotype was lowest for anti-Vi IgG (median 1.5, IQR 1.0 - 2.1) and highest for anti-HlyE IgG (median 30.9, IQR 16.1-54.6) (Figure 11C). Anti-Vi IgG responses 28 days post-infection were elevated 1.5 times above the population mean; anti-HlyE IgG responses were 30 times higher than the population means.

## DISCUSSION

We describe an approach to estimate typhoidal *Salmonella* infection seroincidence from cross-sectional serosurveys using finger-stick dried blood spots. Leveraging longitudinal cohorts of enteric fever cases in four countries, we found IgA and IgG responses to HlyE and LPS were markedly increased for many months following infection, and these elevations were consistent across populations with varying infection pressures. We then used HlyE IgA and IgG to estimate the seroincidence of typhoidal *Salmonella* infection in the catchment areas of six study sites across four countries with prospective blood culture-based surveillance, finding the rank order of seroincidence estimates tracked with clinical enteric fever incidence estimates.

Unlike traditional seroepidemiological methods that rely on a cutoff, we used quantitative, longitudinal antibody responses, integrating IgA and IgG isotype data, to generate population-level seroincidence estimates. A strength of this approach is that we incorporated several types of uncertainty: measurement error, biologic noise, and inter-individual variation in antibody responses. While antibody responses showed considerable individual variation among patients, we found the median fitted peak antibody response was consistent across the four countries. There was some variation in the decay rates across countries, with Bangladesh having the slowest decay rate and Ghana having the fastest. We propose two potential explanations for this difference. The first is that reinfections are more common in settings with a higher force of infection. While we classified suspected reinfections in the longitudinal case data, some reinfections may not have been identified. Second, in settings with a higher force of infection, individuals with typhoid are more likely to be frequently exposed, and secondary antibody responses may wane more slowly.

Previous typhoid seroepidemiological studies have used IgG responses to the Vi polysaccharide capsule. We found the ratio of 28-day antibody responses among cases compared to the population serosurvey participants was lowest for anti-Vi IgG and highest for anti-HlyE IgG, implying anti-Vi IgG has limited utility in distinguishing acute cases from prior exposures. Furthermore, we found no increase in serologic responses to Vi by age, echoing an earlier study from Kathmandu (25). A study from Fiji found anti-Vi IgG responses increase with age but identified no differences in age-response patterns between high and low incidence communities (8). A recent study from Malawi, Nepal, and Bangladesh used a change in Vi IgG response between two longitudinal samples to mark an incident infection and found a similar order of magnitude to the seroincidence estimates presented here (26). However, the use of Vi in WHO-recommended vaccines will preclude distinguishing natural infections from vaccine-derived immunity. Together, these circumstances underscore the importance of using alternative antigens in assessing enteric fever seroincidence.

The seroincidence estimates for all catchment areas were substantially higher than population-based clinical incidence estimates, even after adjusting for care-seeking and blood culture sensitivity. A “high” incidence of clinical enteric fever is defined as >100 cases per 100,000 person-years (27). We estimated seroincidence of >4,000 per 100,000 person-years in all study areas, with rate ratios of seroincidence to clinical incidence ranging from 16 to 32. This implies a substantial incidence of asymptomatic or paucisymptomatic infections, as observed with other infections (6,28–31). One benefit of community-based seroincidence estimates is their robustness to differential care-seeking behavior, health-care access, and cultural differences in reporting illness; this may enable less biased estimates of the force of infection in a community.

In South Asia, 10-20% of enteric fever cases are caused by *S*. Paratyphi A(17). We found antibody responses for *S*. Typhi-LPS were lower among *S*. Paratyphi A cases than among *S*. Typhi cases, which we expected given that *S*. Paratyphi A only shares O12 with *S*. Typhi (has O9 and O12. Additionally, we observed elevated anti-LPS antibody responses among patients with iNTS from Ghana. These cross-reactive responses are likely due to the shared O12 antigen, a trisaccharide repeat backbone identical among *Salmonella* groups A, B, and D (16,32). As such, we used HlyE antibody responses, which did not differ between *S*. Typhi and *S*. Paratyphi A, for our primary seroincidence estimates. Our earlier studies have demonstrated high specificity of anti-HlyE antibody responses for typhoid or paratyphoid diagnosis, compared with other bacterial infections (11,33). We are working to identify *S*. Typhi- or *S*. Paratyphi-specific immune responses to enable serovar-specific seroincidence estimation.

The results of this study should be interpreted within the context of several limitations. First, we incorporated age-dependence in antibody responses among enteric fever cases by fitting separate kinetics models to each age stratum. Methods to formally account for age dependence in the longitudinal decay curves require further development. Second, we focused on our serosurveys on individuals under the age of 26. Characterizing longitudinal antibody responses in older individuals would be needed to calculate seroincidence in older ages and would be of value given the consideration for catch-up TCV campaigns in individuals up to 45 years of age. While the clinical incidence of typhoidal salmonella is higher among older ages in the SEAP study sites in Nepal, we observed a stable seroincidence rate across ages. Possible explanations for this observation are that the ratio between seroincidence and clinical infections might differ by age or that seroincidene estimates may be less reliable at older ages due to multiple exposures and differential waning. The seroincidence estimates reflected the age pattern of clinical incidence for the Bangladesh (Dhaka) and Pakistan (KGH) study sites. Third, we assumed the antibody kinetics were similar for all cases, but it is possible that asymptomatic individuals have different antibody kinetics. We found antibody kinetics did not differ between hospitalized and non-hospitalized enteric fever patients, suggesting that antibody responses are not dependent on clinical disease severity. However, we could not determine whether individuals who have asymptomatic infections or who receive care at lower acuity facilities have similar antibody responses. Our method would underestimate seroincidence if the peak antibody responses are lower and the decay more rapid for asymptomatic cases. Fourth, in regions with high transmission intensity, individuals may be frequently re-exposed, and the shape and parameters of antibody kinetics from secondary and tertiary exposures will likely differ from primary infections (34). Longitudinal studies with serial blood culture are needed to accurately describe the kinetics of true re-infections. Fifth, long-term carriage of typhoidal *Salmonella* may alter the longitudinal serologic responses, though anti-HlyE antibody responses were not identified on an immunoscreen among carriers(35). Sixth, while we know typhoid transmission to be seasonal in some environments, our modeling approach assumes a constant force of infection throughout the year and therefore averages the incidence rate across the year. Finally, while serosurveys may provide an efficient means for incidence estimation in resource-constrained settings, they do not obviate the need for blood culture surveillance, which is the only method for monitoring antimicrobial resistance--a serious and growing threat to the effective treatment of typhoid.

The WHO recommends that countries with high typhoid incidence introduce TCVs in national programs, resulting in an urgent need for typhoidal *Salmonella* incidence data. Clinical surveillance for enteric fever is limited to settings with facilities equipped to perform blood cultures; even when available, incidence estimates may be highly sensitive to biases in care-seeking behaviors, antibiotic use, and variable diagnostic sensitivity (24). We describe a serosurveillance approach that efficiently generates population-level typhoidal *Salmonella* incidence estimates. We found that sample sizes of 200 to 400 individuals per age strata were sufficient to consistently estimate incidence depending on the burden of typhoid in the population, with higher burden settings requiring smaller sample sizes. This approach has the potential to expand the geographic scope of typhoid surveillance, generate much-needed subnational data on its burden, and yield incidence estimates that are comparable across geographic regions and time.

## Data Availability

All data produced in the present study are available upon reasonable request to the authors

## Acknowledgments

This study was supported by a grant from Bill and Melinda Gates Foundation (INV-000572). We would like to acknowledge the essential contributions of field and laboratory teams in Bangladesh: Sultana Aflatun Rubana, Raktim Das, Khairun Naher, Kanis Fatema, Shamima Sultana, Masrufa Akhter, Jarin Sultana, Sathi Akter, Kristina Bain, Lima Akter, Shaswati Gain, Rehana Akter, Morium Akter, Aklima Akter, Khandokar Rehana, Rasheda Khan; Nepal: Sudan Maharjan, Lok Raj Bhatt, Natasha Shrestha, Shishir Ranjit, Anil Khanal, Bipin Khadka, Suman Shrestha, Pusp Raj Bhatt; and Pakistan teams: Kosar Riaz, Shazia Maqsood, Hira Asghar, Naik Banu, Afshan Piyar Ali, Hasina Wajid, Khalida Gul, Salima Shah, Samrina Karim, Faisal Hussain; as well as Caryn Bern and Alexander Yu.We are extremely grateful to all the study participants for their interest and valuable time. Finally, we would like to acknowledge the foundational contributions of Jan van Eijkeren to developing the seroincidence models and advancing methods to incorporate biological and measurement noise.

## Author contributions

Conceptualization: KA, JCS, SKS, FQ, SPL, DOG, JRA

Methodology: KA, JCS, PT, SKS, FQ, SPL, DOG, RCC, JRA

Investigation: KA, JCS, SS, SJM, MSIS, SMAS, AS, NA, FNJ, MSK, DT, KV, R, JS, NK, MTY, JI, IFD, YL, NM, MA, SP, ASC, ATL, CF, ETR, AN, AF, ML, VK, IIB, HJJ, AH, SEP, RZ, FM, EO, YA, MO, PT, DOG, SPL, FNQ, SKS, RCC, JRA

Formal Analysis and Visualization: KA, JCS

Resources: AF, ML, VK, AH, SEP, RZ, FM, EO, YA, MO

Data Curation: JCS, ASC, MSK, SIS, SMAS, NA, KV, NK, YL, NM, MA

Funding acquisition: SPL, DOG, FNQ, SKS, RCC, JRA

Project administration: KA, JCS, ASC, KV, RS, DT, SS, SJM, IFD, DOG, SKS, FQ, JRA

Supervision: SS, SKS, DT, SPL, DOG, FQ, RCC, JRA

Writing – original draft: KA, JCS, RCC, JRA

Writing – review & editing: All authors

Access and verified data: KA, JCS

## Data and materials availability

All analysis code is available at: https://github.com/kaiemjoy/TyphoidSeroIncidence.git

## Disclaimer

The findings and conclusions of this report are those of the authors and do not necessarily represent the official position, policies, or views of the US Centers for Disease Control and Prevention/the Agency for Toxic Substances and Disease Registry.

## References and Notes

1. GBD 2017 Typhoid and Paratyphoid Collaborators. The global burden of typhoid and paratyphoid fevers: a systematic analysis for the Global Burden of Disease Study 2017. Lancet Infect Dis. 2019 Apr;19(4):369–81.

2. Antillon M, Saad NJ, Baker S, Pollard AJ, Pitzer VE. The Relationship Between Blood Sample Volume and Diagnostic Sensitivity of Blood Culture for Typhoid and Paratyphoid Fever: A Systematic Review and Meta-Analysis. J Infect Dis. 2018 Nov 10;218(suppl_4):S255–67.

3. Aiemjoy K, Aragie S, Wittberg DM, Tadesse Z, Callahan EK, Gwyn S, et al. Seroprevalence of antibodies against Chlamydia trachomatis and enteropathogens and distance to the nearest water source among young children in the Amhara Region of Ethiopia. PLoS Negl Trop Dis. 2020 Sep;14(9):e0008647.

4. Arnold BF, Martin DL, Juma J, Mkocha H, Ochieng JB, Cooley GM, et al. Enteropathogen antibody dynamics and force of infection among children in low-resource settings. Ferguson NM, Jit M, White M, Leung DT, Azman A, editors. eLife. 2019 Aug 19;8:e45594.

5. Salje H, Cummings DAT, Rodriguez-Barraquer I, Katzelnick LC, Lessler J, Klungthong C, et al. Reconstruction of antibody dynamics and infection histories to evaluate dengue risk. Nature. 2018 May;557(7707):719–23.

6. de Melker HE, Versteegh FGA, Schellekens JFP, Teunis PFM, Kretzschmar M. The incidence of Bordetella pertussis infections estimated in the population from a combination of serological surveys. J Infect. 2006 Aug;53(2):106–13.

7. Kshatri JS, Bhattacharya D, Praharaj I, Mansingh A, Parai D, Kanungo S, et al. Seroprevalence of SARS-CoV-2 in Bhubaneswar, India: findings from three rounds of community surveys. Epidemiology & Infection [Internet]. 2021 ed [cited 2021 Aug 17];149. Available from: https://www.cambridge.org/core/journals/epidemiology-and-infection/article/seroprevalence-of-sarscov2-in-bhubaneswar-india-findings-from-three-rounds-of-community-surveys/29A7905F297A1EFA68749AC3C6383300

8. Watson CH, Baker S, Lau CL, Rawalai K, Taufa M, Coriakula J, et al. A cross-sectional seroepidemiological survey of typhoid fever in Fiji. PLOS Neglected Tropical Diseases. 2017 Jul 20;11(7):e0005786.

9. Charles RC, Liang L, Khanam F, Sayeed MA, Hung C, Leung DT, et al. Immunoproteomic Analysis of Antibody in Lymphocyte Supernatant in Patients with Typhoid Fever in Bangladesh. Clin Vaccine Immunol. 2014 Mar;21(3):280–5.

10. Charles RC, Sheikh A, Krastins B, Harris JB, Bhuiyan MS, LaRocque RC, et al. Characterization of Anti-Salmonella enterica Serotype Typhi Antibody Responses in Bacteremic Bangladeshi Patients by an Immunoaffinity Proteomics-Based Technology. Clin Vaccine Immunol. 2010 Aug;17(8):1188–95.

11. Andrews JR, Khanam F, Rahman N, Hossain M, Bogoch II, Vaidya K, et al. Plasma Immunoglobulin A Responses Against 2 Salmonella Typhi Antigens Identify Patients With Typhoid Fever. Clin Infect Dis. 2019 Mar 5;68(6):949–55.

12. Faucher SP, Forest C, Béland M, Daigle F. A novel PhoP-regulated locus encoding the cytolysin ClyA and the secreted invasin TaiA of Salmonella enterica serovar Typhi is involved in virulence. Microbiology (Reading). 2009 Feb;155(Pt 2):477–88.

13. Fuentes JA, Villagra N, Castillo-Ruiz M, Mora GC. The Salmonella Typhi hlyE gene plays a role in invasion of cultured epithelial cells and its functional transfer to S. Typhimurium promotes deep organ infection in mice. Res Microbiol. 2008 May;159(4):279–87.

14. Oscarsson J, Westermark M, Löfdahl S, Olsen B, Palmgren H, Mizunoe Y, et al. Characterization of a Pore-Forming Cytotoxin Expressed by Salmonella enterica Serovars Typhi and Paratyphi A. Infect Immun. 2002 Oct;70(10):5759–69.

15. McClelland M, Sanderson KE, Clifton SW, Latreille P, Porwollik S, Sabo A, et al. Comparison of genome degradation in Paratyphi A and Typhi, human-restricted serovars of Salmonella enterica that cause typhoid. Nat Genet. 2004 Dec;36(12):1268–74.

16. Liu D, Verma NK, Romana LK, Reeves PR. Relationships among the rfb regions of Salmonella serovars A, B, and D. J Bacteriol. 1991 Aug;173(15):4814–9.

17. Garrett D, Longley A, Aiemjoy K, Qamar FN, Saha SK, Yousafzai MT, et al. Incidence of Typhoid and Paratyphoid Fever in Bangladesh, Nepal, and Pakistan: Results of the Surveillance for Enteric Fever in Asia Project. Lancet Global Health [Internet]. 2022 Mar 8 [cited 2022 Mar 8];In Press. Available from: https://papers.ssrn.com/abstract=3866551

18. Park SE, Toy T, Cruz Espinoza LM, Panzner U, Mogeni OD, Im J, et al. The Severe Typhoid Fever in Africa Program: Study Design and Methodology to Assess Disease Severity, Host Immunity, and Carriage Associated With Invasive Salmonellosis. Clinical Infectious Diseases. 2019 Oct 30;69(Supplement_6):S422–34.

19. de Graaf WF, Kretzschmar MEE, Teunis PFM, Diekmann O. A two-phase within-host model for immune response and its application to serological profiles of pertussis. Epidemics. 2014 Dec;9:1–7.

20. Teunis PFM, van Eijkeren JCH, de Graaf WF, Marinović AB, Kretzschmar MEE. Linking the seroresponse to infection to within-host heterogeneity in antibody production. Epidemics. 2016 Sep;16:33–9.

21. Plummer M, Stukalov A, Denwood M, Plummer MM. Package ‘rjags.’ 2019.

22. Teunis PFM, van Eijkeren JCH. Estimation of seroconversion rates for infectious diseases: Effects of age and noise. Stat Med. 2020 Sep 20;39(21):2799–814.

23. Teunis PFM, van Eijkeren JCH, Ang CW, van Duynhoven YTHP, Simonsen JB, Strid MA, et al. Biomarker dynamics: estimating infection rates from serological data. Stat Med. 2012 Sep 10;31(20):2240–8.

24. Andrews JR, Barkume C, Yu AT, Saha SK, Qamar FN, Garrett D, et al. Integrating Facility-Based Surveillance With Healthcare Utilization Surveys to Estimate Enteric Fever Incidence: Methods and Challenges. J Infect Dis. 2018 Nov 10;218(suppl_4):S268–76.

25. Pulickal AS, Gautam S, Clutterbuck EA, Thorson S, Basynat B, Adhikari N, et al. Kinetics of the Natural, Humoral Immune Response to Salmonella enterica Serovar Typhi in Kathmandu, Nepal. Clinical and Vaccine Immunology. 2009 Oct 1;16(10):1413–9.

26. Meiring JE, Shakya M, Khanam F, Voysey M, Phillips MT, Tonks S, et al. Burden of enteric fever at three urban sites in Africa and Asia: a multicentre population-based study. The Lancet Global Health. 2021 Dec 1;9(12):e1688–96.

27. Crump JA, Luby SP, Mintz ED. The global burden of typhoid fever. Bull World Health Organ. 2004 May;82(5):346–53.

28. Monge S, Teunis P, Friesema I, Franz E, Ang W, Pelt W van, et al. Immune response-eliciting exposure to Campylobacter vastly exceeds the incidence of clinically overt campylobacteriosis but is associated with similar risk factors: A nationwide serosurvey in the Netherlands. Journal of Infection. 2018 Sep 1;77(3):171–7.

29. Simonsen J, Mølbak K, Falkenhorst G, Krogfelt KA, Linneberg A, Teunis PFM. Estimation of incidences of infectious diseases based on antibody measurements. Statistics in Medicine. 2009;28(14):1882–95.

30. Byambasuren O, Dobler CC, Bell K, Rojas DP, Clark J, McLaws M-L, et al. Comparison of seroprevalence of SARS-CoV-2 infections with cumulative and imputed COVID-19 cases: Systematic review. PLOS ONE. 2021 Apr 2;16(4):e0248946.

31. Bobrovitz N, Arora RK, Cao C, Boucher E, Liu M, Donnici C, et al. Global seroprevalence of SARS-CoV-2 antibodies: A systematic review and meta-analysis. PLOS ONE. 2021 Jun 23;16(6):e0252617.

32. Wahid R, Simon R, Zafar SJ, Levine MM, Sztein MB. Live oral typhoid vaccine Ty21a induces cross-reactive humoral immune responses against Salmonella enterica serovar Paratyphi A and S. Paratyphi B in humans. Clin Vaccine Immunol. 2012 Jun;19(6):825–34.

33. Kumar S, Nodoushani A, Khanam F, DeCruz AT, Lambotte P, Scott R, et al. Evaluation of a Rapid Point-of-Care Multiplex Immunochromatographic Assay for the Diagnosis of Enteric Fever. mSphere. 2020 Jun 10;5(3).

34. Diekmann O, de Graaf WF, Kretzschmar MEE, Teunis PFM. Waning and boosting: on the dynamics of immune status. J Math Biol. 2018 Dec;77(6–7):2023–48.

35. Charles RC, Sultana T, Alam MM, Yu Y, Wu-Freeman Y, Bufano MK, et al. Identification of Immunogenic Salmonella enterica Serotype Typhi Antigens Expressed in Chronic Biliary Carriers of S. Typhi in Kathmandu, Nepal. PLOS Neglected Tropical Diseases. 2013 Aug 1;7(8):e2335.

