## Appendix for "Estimating typhoid incidence from community-based serosurveys: A multicohort study in Bangladesh, Nepal, Pakistan and Ghana"

### Appendix to: 'Novel seroepidemiological approaches for estimating typhoid incidence: a multicohort study in Bangladesh, Nepal, Pakistan and Ghana'

TABLE OF CONTENTS


I. **SUPPLEMENTAL METHODS**
   A. Laboratory methods
   B. Statistical methods
      1. Reinfections among blood culture-confirmed cases
      2. Measurement error, biologic noise and lower limit of detection parameters for incidence estimation
      3. Sample size
   C. References

II. **SUPPLEMENTAL FIGURES**
   A. Fig. S1. Time of sample collection following fever onset among culture-confirmed enteric fever cases by age and country.
   B. Fig. S2. Anti-HlyE and LPS IgG and IgA responses among cases, population serosurvey participants and North American controls.
   C. Fig. S3. Intraindividual correlation in anti-HlyE and anti-LPS IgG and IgA responses at 1, 6, and 11+ months after fever onset in cases, by age category.
   D. Fig. S4. Kinetics of anti-HlyE and anti-LPS IgG and IgA responses among culture-confirmed enteric fever cases by hospitalization status.
   E. Fig. S5. Study flow diagram for population-based serosurvey enrollment.
   F. Fig. S6. Comparison of age-specific seroincidence estimates using overall and country-specific longitudinal parameters.
   G. Fig. S7. Anti-HlyE and anti-LPS IgG and IgA responses at enrollment with acute febrile illness, among participants with culture-confirmed invasive non-typhoidal *Salmonellosis*, *S.* Typhi, and *S.* Paratyphi A infections.
   H. Fig. S8. Relationship between serosurvey size and estimated seroincidence for a high (Dhaka) and more moderate (Kavre) incidence community.

III. **SUPPLEMENTAL TABLES**
   A. Table S1: Model parameter estimates for antibody kinetics by antigen, immunoglobulin isotype and age group.
   B. Table S2: Estimates of typhoidal *Salmonella* infection from cross-sectional serological data by study community, age group, and antigen/isotype combinations used in model.
   C. Table S3: Measurement error, biologic noise and lower censoring limits for each antigen-isotype

#### I. SUPPLEMENTAL METHODS

##### Laboratory methods

We cut two filled, dried blood spot (DBS) filter paper protrusions and submerged them in 133 $\mu$L of 1XPBS 0.05% Tween buffer overnight at 4℃; and eluates were recovered after centrifugation. The eluate was used immediately or stored at 4℃ for up to 2 weeks. We coated plates with *S.* Typhi LPS (2.5 µg/mL) purified from strain Ty21a (2.5 µg/mL), purified HlyE (1 µg/mL) or Vi antigen (Sanofi Pasteur; 2 µg/mL) as previously described (1,2). Plasma was added to the plate in duplicate at the following dilutions for each antigen and antibody isotype (IgA, IgG, respectively): LPS (1:1000; 1:5000), HlyE (1:500, 1:5000), Vi (1:100 IgG only, Nepal and Ghana only). DBS eluate was assumed to be 1:10 dilution of plasma, which accounts for the dilution of plasma during the DBS elution. Bound antibodies were detected with goat anti-human IgG and IgA conjugated with horseradish peroxidase (Jackson ImmunoResearch), and peroxidase activity was measured at 450 nm using the chromogenic substrate, o-phenylenediamine. To compare across ELISA plates and sites, the blank-adjusted sample readings were averaged, divided by the readings of a standard included on each plate (plasma pool for LPS and Vi; human chimeric monoclonal antibody for HlyE), multiplied by 100 and reported as ELISA units.

##### Statistical methods

**Reinfections among blood culture-confirmed cases:** We defined suspected reinfections as cases with a 3-times or higher increase for 2 or more antigen-isotypes occurring at least 3 months after their initial infection episode. The 3-times threshold was derived by calculating the median times change from baseline to 28 days among blood culture-confirmed cases. For antibody decay estimates, we excluded longitudinal observations from individuals who met the definition of a suspected reinfection from the time of the identified reinfection and all subsequent remaining follow-up observations. We calculated the incidence of suspected reinfection by dividing the total number of possible reinfections identified by the sum of the observed person-time in each country; we used bootstrap resampling to calculate 95% confidence intervals.

**Measurement error, biologic noise and lower limit of detection parameters for incidence estimation:** We accounted for two sources of noise in the serologic responses: measurement noise of the assay (described by the coefficient of variation [CV] across replicates) and biologic noise (measured as background response to the antigen-isotype among never-exposed, negative controls) as detailed in (3). We calculated the coefficients of variation (CV) from a panel of 100 negative and positive controls that were run in triplicate at each site. We estimated biologic noise by running the ELISAs on used stored serum samples from children enrolled in two studies on celiac disease: 1) 48 children (1-5 years) who had first degree relatives with celiac disease, enrolled nationally and 2) 31 healthy controls (2-18 years), enrolled at Massachusetts General Hospital (MGH, Boston, Massachusetts, USA) (4). To calculate lower detection limits we conducted serial dilutions of each antigen isotype and chose a threshold at which the CV was greater than or equal to 30% and added two standard deviations (5) then normalized to the positive control. Lower detection limits were estimated for each laboratory running the assays. Measurement and biological noise parameters and the limits of detection used for each site are detailed in Table S3.

**Sample size:** To determine a sufficient sample size to consistently characterize incidence, we resampled the catchment area population serosurvey data and estimated the seroincidence using various sample sizes ranging from 50 to 600. We used bootstrapping to select 100 samples with replacement for each sample size and estimated the incidence in each sample. We then calculated the percent of resampled estimates falling within the 95% confidence intervals for the full-serosurvey estimate and the half-width of the confidence intervals relative to the incidence estimates.

With sample sizes of 100 individuals per serosurvey, the percent of bootstrapped re-sampled incidence estimates falling within the 95% CI of the full estimate (ie coverage) was 88% in Dhaka and 72% in Kavre; With a sample size of 300, coverage increased to 100% in Dhaka and 87% in Kavre and with 600 coverage for Kavre reached 95% with a sample size of 300 and 100% in Dhaka and 95% in Kavre with a sample size of 600 (Figure S8A). The average half-width of the confidence interval relative to the incidence estimate fell below 25% at a sample size of 250 for Dhaka and 450 for Kavre (Figure S8B).

**References**

1. Andrews JR, Khanam F, Rahman N, Hossain M, Bogoch II, Vaidya K, et al. Plasma Immunoglobulin A Responses Against 2 Salmonella Typhi Antigens Identify Patients With Typhoid Fever. Clin Infect Dis. 2019 Mar 5;68(6):949–55.
2. Charles RC, Sultana T, Alam MM, Yu Y, Wu-Freeman Y, Bufano MK, et al. Identification of immunogenic Salmonella enterica serotype Typhi antigens expressed in chronic biliary carriers of S. Typhi in Kathmandu, Nepal. PLoS Negl Trop Dis. 2013;7(8):e2335.
3. Teunis PFM, van Eijkeren JCH. Estimation of seroconversion rates for infectious diseases: Effects of age and noise. Stat Med. 2020 Sep 20;39(21):2799–814.
4. Leonard MM, Camhi S, Huedo-Medina TB, Fasano A. Celiac Disease Genomic, Environmental, Microbiome, and Metabolomic (CDGEMM) Study Design: Approach to the Future of Personalized Prevention of Celiac Disease. Nutrients. 2015 Nov 11;7(11):9325–36.
5. EP17A2 | Evaluation of Detection Capability for Clinical Laboratory Measurement Procedures, 2nd Edition [Internet]. Clinical & Laboratory Standards Institute. [cited 2021 Sep 10]. Available from: https://clsi.org/standards/products/method-evaluation/documents/ep17/

#### II. SUPPLEMENTAL FIGURES

**Fig S1. Time of sample collection in the population-based serosurveys.** Each point represents an individual study participant.

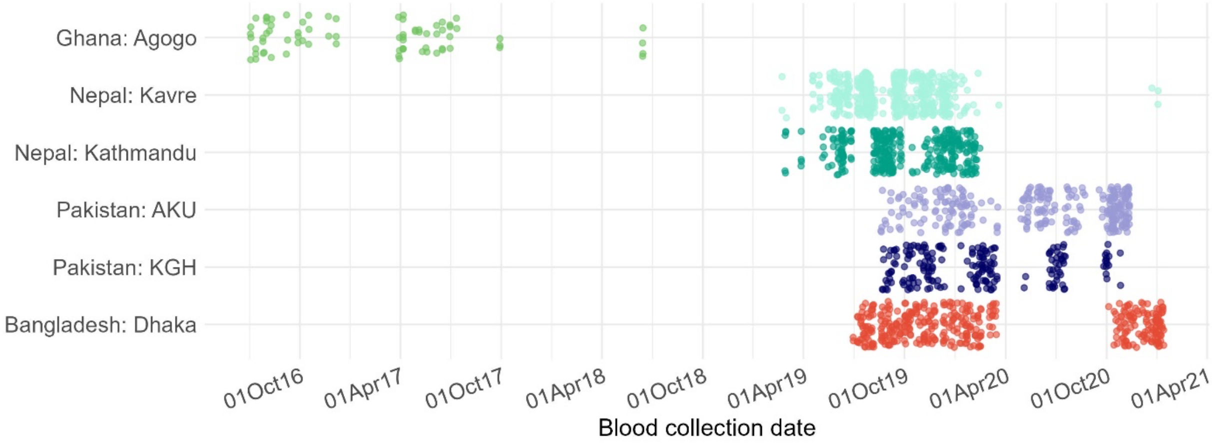

**Fig S2. Correlation between Anti-HlyE and anti-LPS IgG and IgA responses measured from DBS and Plasma.** Each dot indicates an individual's quantitative antibody response measured in ELISA units.

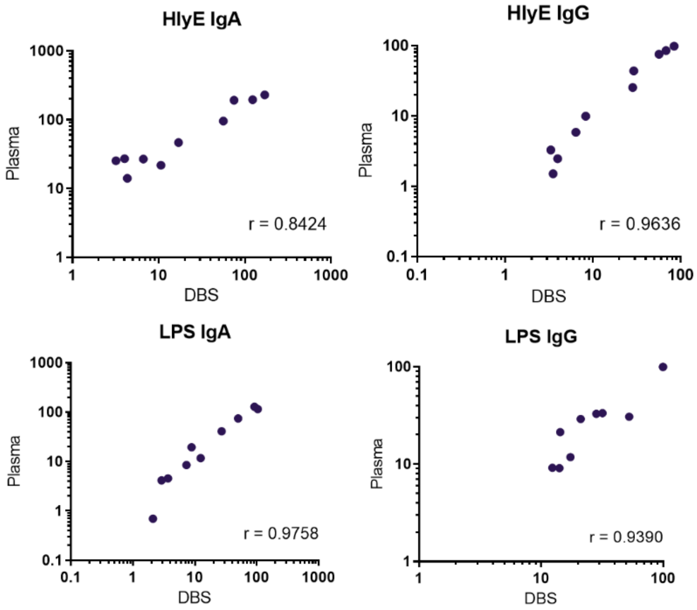

**Fig. S3. Time of sample collection following fever onset among culture-confirmed enteric fever cases by age and country.** Each line represents an individual study participant and each point represents a time point of blood sample collection. The age distribution of the cases reflects the case burden as captured in the SEAP and SETA studies.

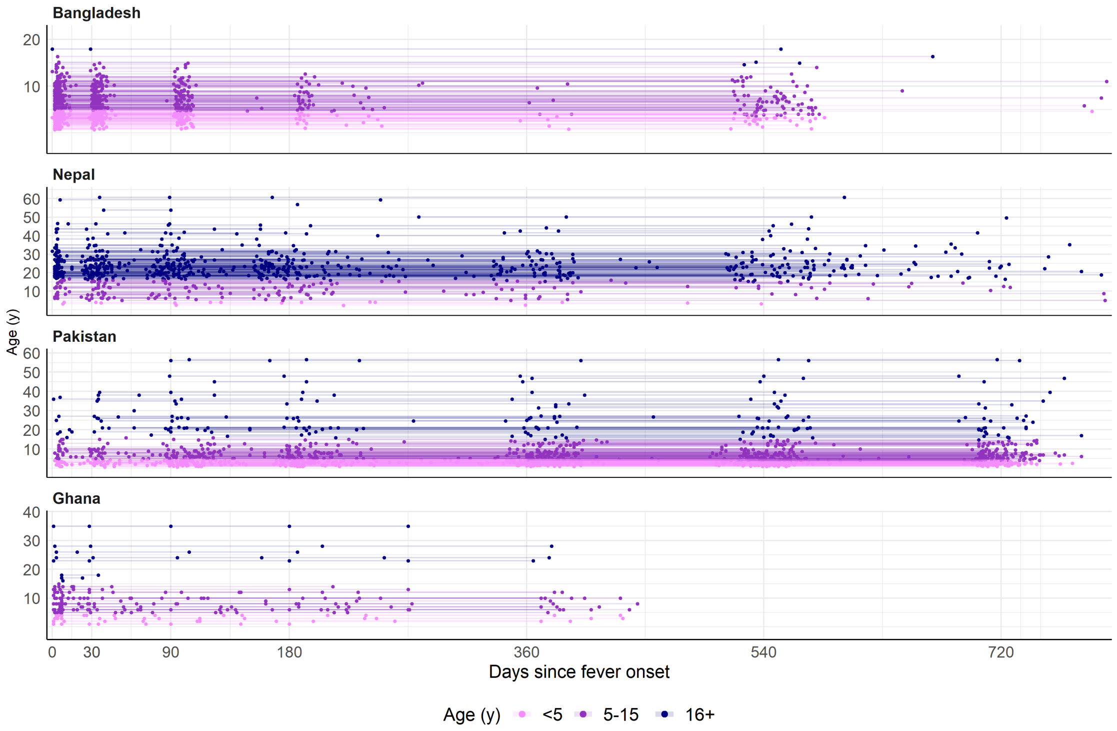

**Fig. S4. Anti-HlyE and LPS IgG and IgA responses among cases, population serosurvey participants and North American controls.**

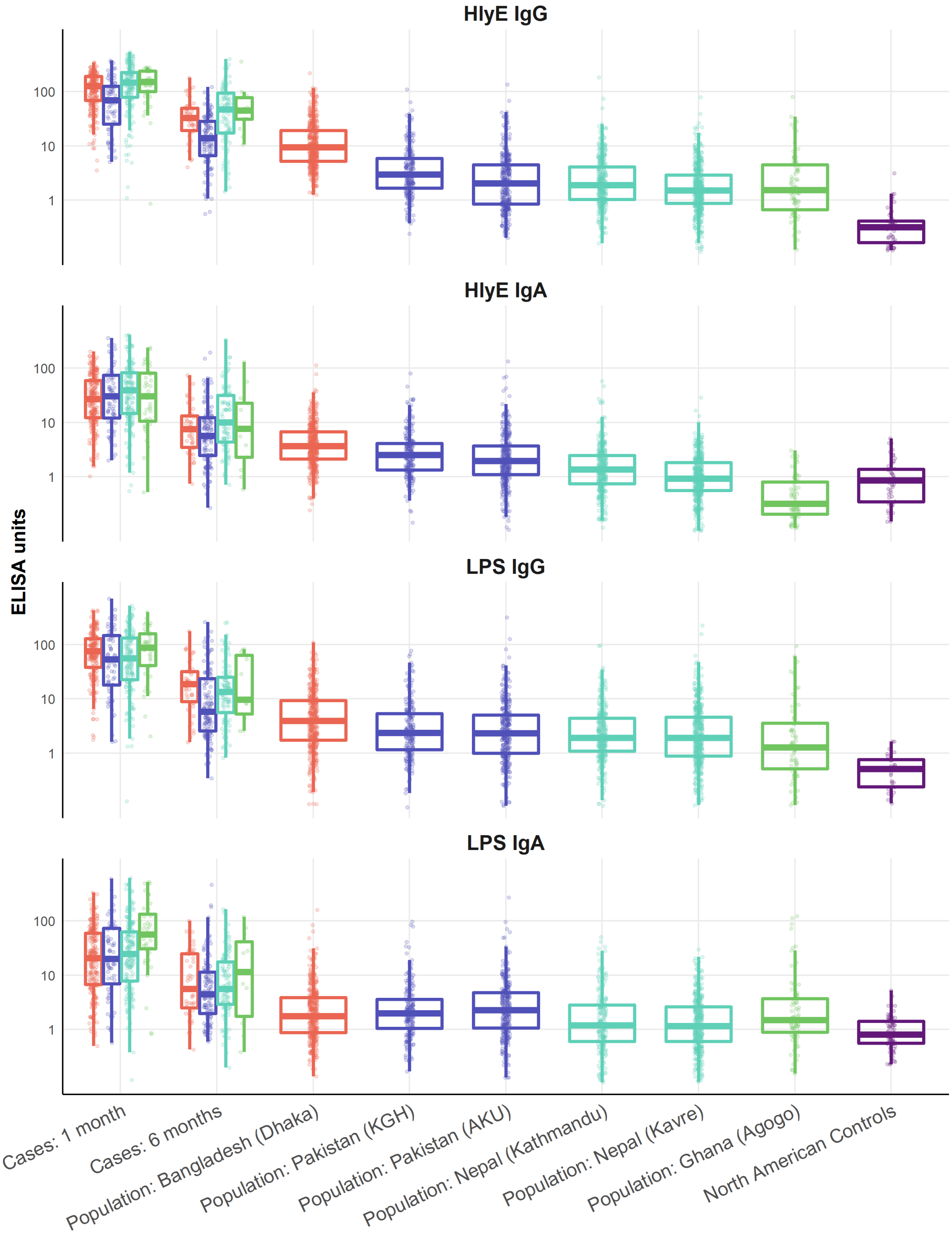

**Fig. S5. Intraindividual correlation in anti-HlyE and anti-LPS IgG and IgA responses at 1, 6, and 11+ months after fever onset in cases, by age category.** Numbers reflect Spearman correlations of paired measurements within individuals, with darker box colors indicating higher correlation.

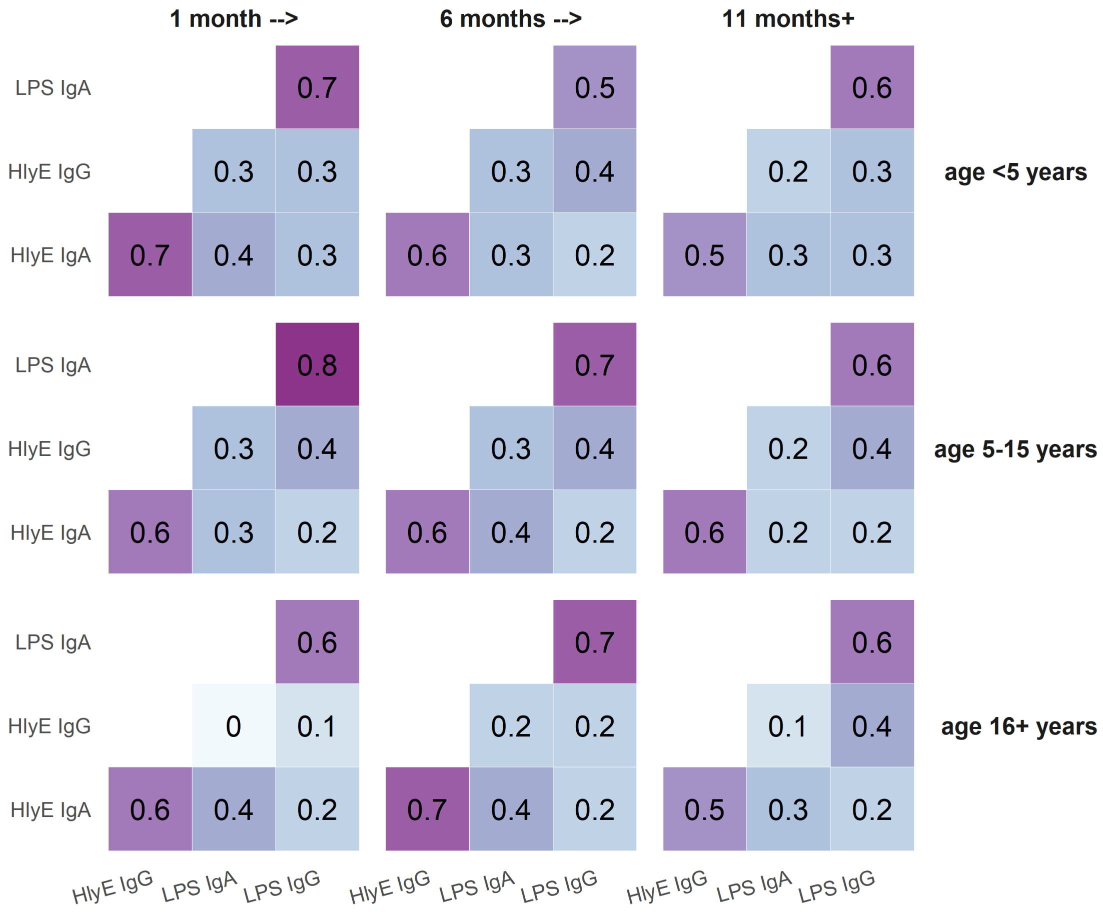

**Fig. S6. Kinetics of anti-HlyE and anti-LPS IgG and IgA responses among culture-confirmed enteric fever cases by hospitalization status.** Comparison was restricted to participants aged 5 to 15 years and includes cases from all four countries. The light-colored lines are the observed individual antibody concentrations; each line indicates one patient. The dark solid and dotted lines indicate the median and 95% credible intervals for the model-fitted antibody decay concentrations.

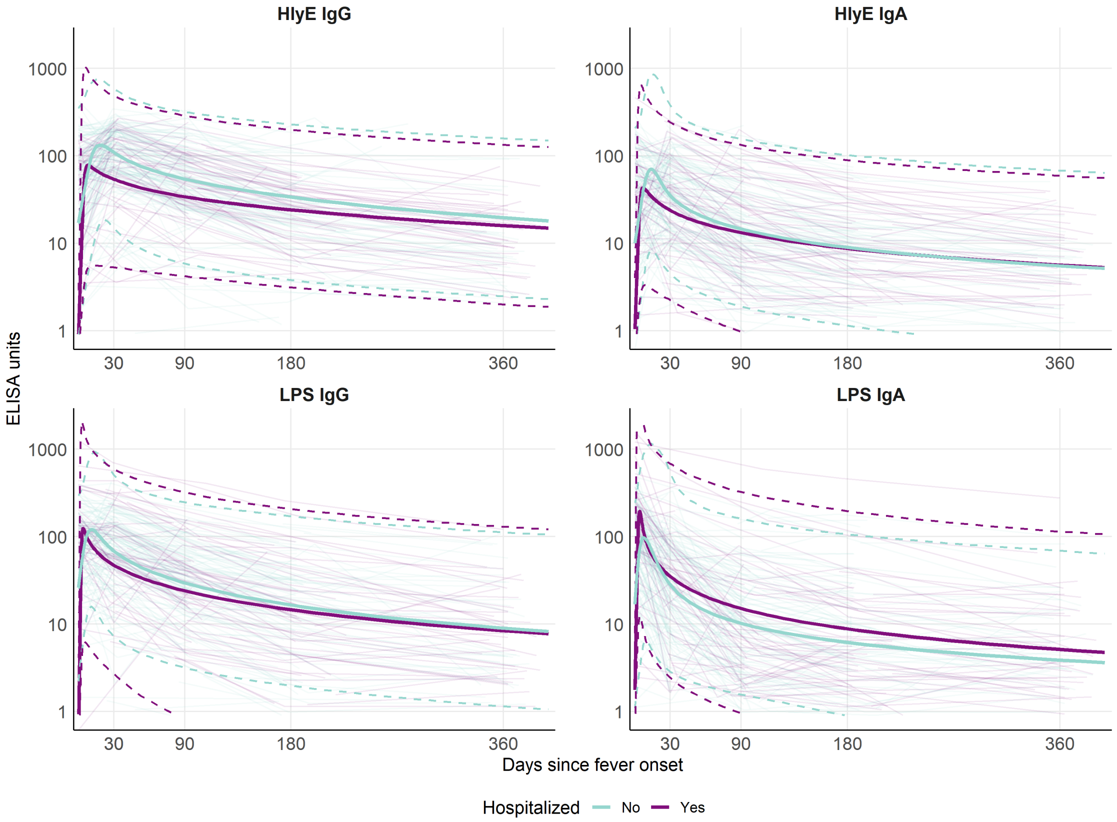


**Fig. S7. Study flow diagram for population-based serosurvey enrollment.**

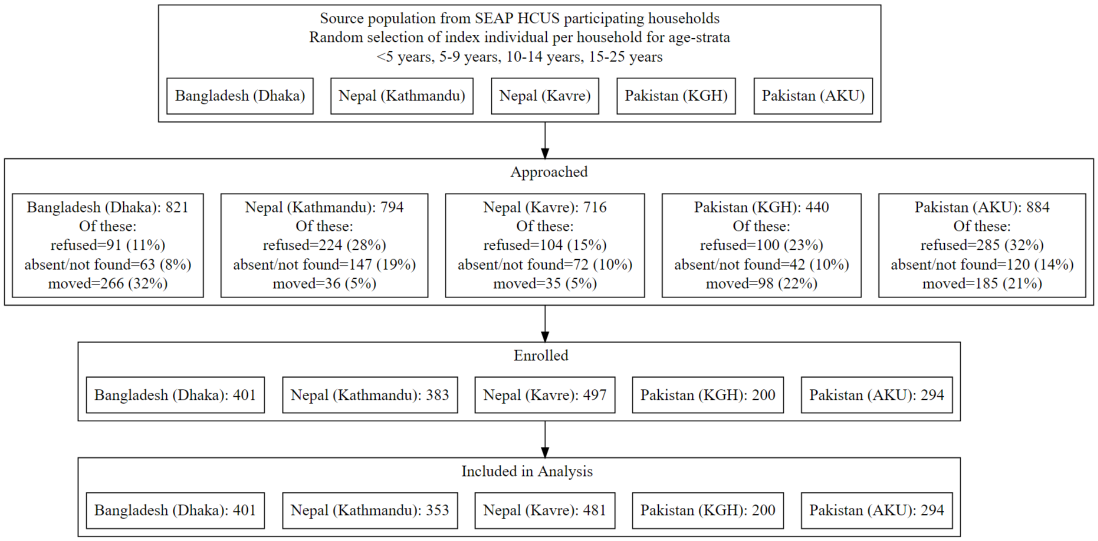

**Fig. S8. Comparison of age-specific seroincidence estimates using overall and country-specific longitudinal parameters.** Seroincidence estimates using overall longitudinal parameters (in light purple) and country-specific longitudinal parameters (in dark purple) for each age group. No country-specific estimates could be generated for the Nepal sites age <5 or the Bangladesh site age 16+ because there were insufficient cases in these sub-populations to support stratified longitudinal kinetic models.

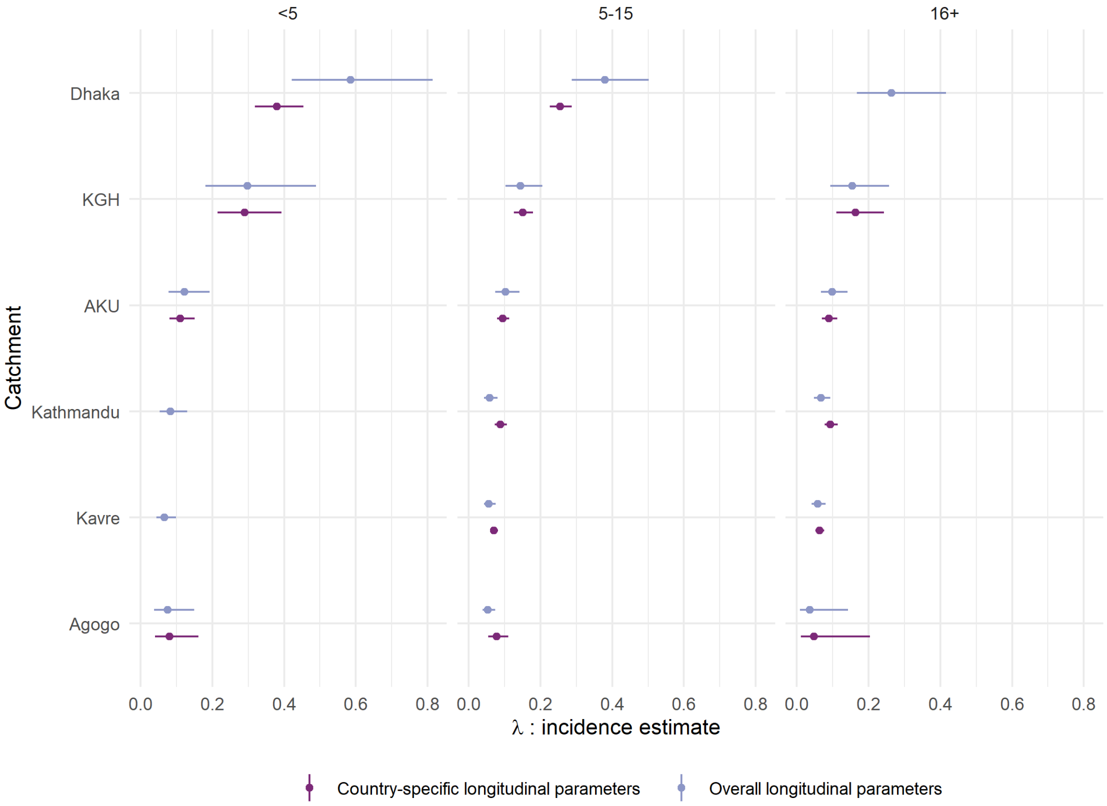

**Fig. S9. Anti-HlyE and anti-LPS IgG and IgA responses at enrollment with acute febrile illness, among participants with culture-confirmed invasive non-typhoidal *Salmonellosis*, *S.* Typhi, and *S.* Paratyphi A infections.** Each dot indicates the quantitative response on a case measured in ELISA units, with boxplot overlaid. All iNTS cases were from the Ghana study site. Baseline is defined as the initial study visit and is restricted to samples collected within 7 days of fever onset.

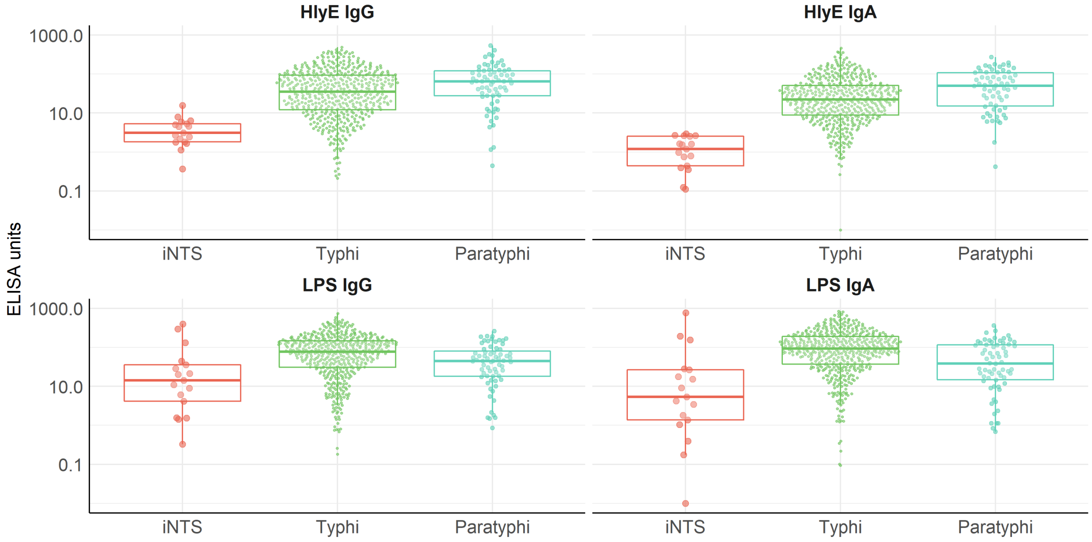

**Fig. S10. Relationship between serosurvey size and estimated seroincidence for a high (Dhaka) and more moderate (Kavre) incidence community.** 100 samples were drawn with sizes ranging from 50 to 600 from each catchment area population. (A) The vertical black dotted line and grey shaded area indicates the seroincidence rate and 95% Confidence Intervals (CI) estimated from the full population-based serosurvey. Each horizontal line represents the seroincidence rate and 95% CI estimated for a re-sampled population. The horizontal lines are colored according to whether the incidence estimate fell within (in purple) or outside (in light blue) the confidence interval of the full serosurvey estimate. The numeric proportion of incidence estimates that fell within the 95% CIs of the full serosurvey estimate are shown in purple in the bottom right corner of each box. (B) Each dot represents the half-width of the confidence interval estimated from a re-sampled population at each sample size.

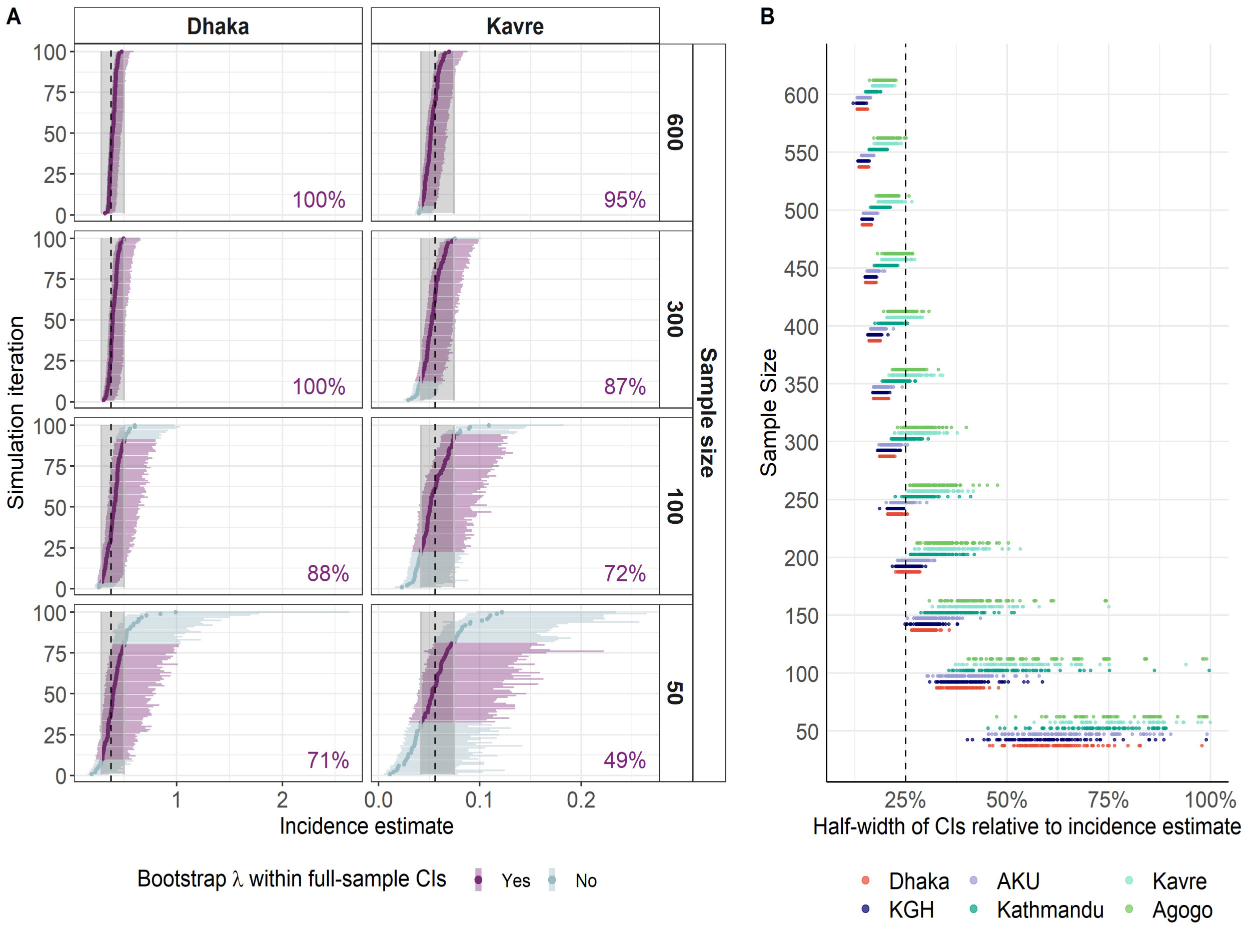

#### III. SUPPLEMENTAL TABLES

**Table S1** Model parameter estimates for antibody kinetics by antigen, immunoglobulin isotype and age group.

| | HlyE IgA | HlyE IgG | LPS IgA | LPS IgG | Vi IgG |
|---|---|---|---|---|---|
| **Peak antibody response (y1)** | | | | | |
| <5 | 37.55 (7-469.8) | 159.67 (20.7-1686.8) | 81.66 (25.8-254.6) | 157.12 (23.3-1601.8) | 325.44 (7.4-22290.5) |
| 5-15 | 103.32 (13-1385.9) | 222.71 (49.6-1541.7) | 338.61 (56.7-8402.8) | 219.99 (48.9-4235.2) | 399.55 (147.4-6077.6) |
| 16+ | 130.92 (10.1-2569.6) | 248.62 (30-2726.3) | 88.73 (4-2788.1) | 112.84 (8.9-1625.4) | 650.91 (218.5-3107.3) |
| Overall | 93.98 (14.1-892.2) | 226.26 (56.5-1096.1) | 271.06 (31.3-3949.3) | 200.54 (47.9-1550) | 542.7 (143.5-3015.6) |
| **Decay Rate (α)** | | | | | |
| <5 | 0.03 (0-1.5) | 0.04 (0-0.6) | 0.05 (0-4.4) | 0.11 (0-1.5) | 0 (0-0) |
| 5-15 | 0.03 (0-2.3) | 0.03 (0-0.8) | 0.06 (0-4.3) | 0.08 (0-1.6) | 0.02 (0-3.9) |
| 16+ | 0.02 (0-0.5) | 0.04 (0-0.3) | 0.01 (0-1.9) | 0.05 (0-0.5) | 0.01 (0-0.1) |
| Overall | 0.03 (0-1.8) | 0.02 (0-0.6) | 0.03 (0-3.6) | 0.07 (0-1.3) | 0 (0-0.4) |
| **Shape factor (r)** | | | | | |
| <5 | 3.24 (1.8-7) | 2.2 (1.6-3.2) | 3.26 (1.6-10.3) | 2.19 (1.3-5.1) | 2.5 (1.2-4.7) |
| 5-15 | 2.58 (1.7-4.5) | 2.08 (1.4-4) | 2.48 (1.3-8.6) | 2.07 (1.4-3.6) | 1.28 (1-2.8) |
| 16+ | 2.64 (1.9-4.1) | 2.01 (1.4-3.4) | 3.26 (2.4-4.9) | 2.26 (1.8-3) | 1.35 (1.2-1.8) |
| Overall | 2.81 (1.7-5.7) | 2.18 (1.5-3.9) | 2.89 (1.5-7.9) | 2.16 (1.4-4.1) | 1.25 (1-3.1) |
| **Time to peak antibody response (t1)** | | | | | |
| <5 | 28.53 (8-85.1) | 16.15 (2.3-104.5) | 9.3 (2.3-33.7) | 6.2 (0.6-56.3) | 1.4 (0.6-3.3) |
| 5-15 | 19.95 (6.4-54) | 12.34 (3.5-41.4) | 6.72 (1.2-32.4) | 6.19 (1.1-32.9) | 4.71 (2.1-10.4) |
| 16+ | 20.76 (6-66.4) | 19.06 (6.4-46.8) | 26.52 (13.7-52.1) | 10.74 (1.9-43.2) | 9.57 (2.8-25.7) |
| Overall | 19.63 (5.2-70.8) | 15.6 (3.8-63.7) | 11.07 (2.5-36.9) | 5.48 (0.7-37.1) | 2.92 (0.6-11.3) |
| **Baseline antibody response (y0)** | | | | | |
| <5 | 20.77 (2.4-181.4) | 14.67 (1.2-181.2) | 77.47 (24-216.8) | 10.6 (0.5-228.8) | 0.6 (0.2-1.7) |
| 5-15 | 14.93 (0.7-295.4) | 15.06 (0.7-313.2) | 19.67 (1.1-298.6) | 20.78 (1.2-344.1) | 164.55 (64.2-354.4) |
| 16+ | 13.99 (0.7-290.1) | 15.59 (0.7-381.4) | 68.8 (3.2-1441.8) | 32.72 (1.8-539.2) | 201.78 (22.3-1762.1) |
| Overall | 13.81 (0.8-254.5) | 14.47 (0.6-323.9) | 32.67 (1.4-655.9) | 13.31 (0.5-302.1) | 126.72 (7.6-1806.3) |

**Table S2** Estimates of typhoidal *Salmonella* seroincidence from cross-sectional serological data by study community, age group, and antigen/isotype combinations used in model.

| | Typhoidal Salmonella seroincidence rate per 100 person-years | |
|---|---|---|
| **Age strata** | **HlyE IgG & IgA** | **HlyE & LPS IgG & IgA** |
| **Bangladesh: Dhaka** | | |
| <5 | 58.5 (42.1-81.4) | 33.8 (24.9-45.9) |
| 5-15 | 37.9 (28.7-50.2) | 22.2 (17.4-28.5) |
| 16+ | 26.3 (16.7-41.5) | 17.0 (11.1-25.9) |
| Overall | 41.2 (34.0-50.1) | 24.3 (20.4-29.0) |
| **Pakistan: KGH** | | |
| <5 | 29.7 (18.1-48.9) | 23.4 (14.5-37.7) |
| 5-15 | 14.5 (10.2-20.6) | 12.6 (9.0-17.6) |
| 16+ | 15.4 (9.2-25.7) | 12.7 (7.8-20.7) |
| Overall | 17.6 (13.7-22.6) | 14.7 (11.6-18.7) |
| **Pakistan: AKU** | | |
| <5 | 12.2 (7.7-19.2) | 12.4 (8.3-18.5) |
| 5-15 | 10.2 (7.4-14.1) | 11.1 (8.3-14.8) |
| 16+ | 9.8 (6.8-14.1) | 8.8 (6.2-12.5) |
| Overall | 10.5 (8.5-13.0) | 10.6 (8.7-12.9) |
| **Nepal: Kathmandu** | | |
| <5 | 8.3 (5.4-12.9) | 6.1 (3.9-9.4) |
| 5-15 | 5.8 (4.2-8.0) | 4.8 (3.5-6.5) |
| 16+ | 6.6 (4.7-9.3) | 6.5 (4.8-9.0) |
| Overall | 6.6 (5.4-8.1) | 5.7 (4.6-6.9) |
| **Nepal: Kavre** | | |
| <5 | 6.6 (4.3-9.9) | 5.1 (3.4-7.6) |
| 5-15 | 5.6 (4.2-7.4) | 4.6 (3.5-6.1) |
| 16+ | 5.8 (4.2-8.0) | 5.8 (4.3-7.9) |
| Overall | 5.8 (4.8-7.1) | 5.1 (4.3-6.1) |
| **Ghana: Agogo** | | |
| <5 | 7.5 (3.8-15.0) | 9.3 (5.7-15.0) |
| 5-15 | 5.3 (3.8-7.4) | 6.9 (5.5-8.6) |
| 16+ | 3.6 (0.9-14.2) | 4.0 (1.5-11.1) |
| Overall | 5.5 (4.1-7.3) | 6.9 (5.7-8.4) |

**Table S3** Measurement error, biologic noise and lower censoring limits for each antigen-isotype.

| Country | Biologic noise* | Measurement error (CV) | Lower censoring limit |
|---|---|---|---|
| **HlyE IgA** | | | |
| Bangladesh | 2.87 | 0.28 | 0.38 |
| Ghana | 2.87 | 0.24 | 0.18 |
| Nepal | 2.87 | 0.24 | 0.85 |
| Pakistan | 2.87 | 0.28 | 0.51 |
| **HlyE IgG** | | | |
| Bangladesh | 1.08 | 0.31 | 0.79 |
| Ghana | 1.08 | 0.16 | 0.65 |
| Nepal | 1.08 | 0.13 | 1.89 |
| Pakistan | 1.08 | 0.15 | 1.59 |
| **LPS IgA** | | | |
| Bangladesh | 2.72 | 0.30 | 0.66 |
| Ghana | 2.72 | 0.16 | 0.86 |
| Nepal | 2.72 | 0.11 | 1.79 |
| Pakistan | 2.72 | 0.25 | 5.13 |
| **LPS IgA** | | | |
| Bangladesh | 0.96 | 0.30 | 0.99 |
| Ghana | 0.96 | 0.20 | 0.88 |
| Nepal | 0.96 | 0.18 | 0.65 |
| Pakistan | 0.96 | 0.27 | 4.84 |

* upper 95% from North American controls

† All values are on the natural scale (not log transformed)
